# Metabolomic breath landscape analysis unravels lipid biomarker candidates in patients with monogenic and idiopathic Parkinson’s disease

**DOI:** 10.1101/2025.04.17.25326025

**Authors:** Madiha Malik, Norbert Brüggemann, Tatiana Usnich, Max Borsche, Tobias Demetrowitsch, Karin Schwarz, Peter Bauer, Katja Lohmann, Christine Klein, Thomas Kunze

## Abstract

Parkinson’s disease (PD) is the fastest growing neurodegenerative disorder. Lack of efficient early diagnostic tools highlights the critical necessity for novel approaches in biomarker discovery. We propose an untargeted metabolomics approach using non-invasive exhaled breath analysis. Breath samples, collected from 73 PD patients, encompassing both genetic (*LRRK2*: n=12, *GBA1*: n=35, *PRKN*: n=6) and idiopathic PD (n=20), 4 unaffected *LRRK2* carriers and 90 controls underwent extreme-resolution FT-ICR-MS analysis. Findings were compared with metabolomics data from blood plasma. Biostatistical analyses identified discernible metabolic patterns in both biofluids, enabling differentiation of PD patients from healthy controls (OOB error < 1%). Metabolomic breath profiling of PD patients yielded 10 significant hits putatively identified as tetracosanoic acid, tricosanoic acid, HMVA, docosanamide, eicosanoic acid, nonadecanoic acid, homophytanic acid, nonadecyl-MG, stearic acid and palmitic acid in PD patients, irrespective of the genetic status. Most of the proposed structures are intermediates in fatty acid metabolism, introducing new candidate biomarkers for breath analysis in PD. Seven of these metabolites were also found in unaffected carriers of pathogenic variants in *LRRK2* when compared to controls. Breath analysis effectively distinguishes between PD patients and healthy controls and nominates metabolites that could serve as noninvasive biomarkers for PD, potentially including its presymptomatic stage.

## INTRODUCTION

Over the past decade, Parkinson’s disease (PD) has shown the most rapid growth among neurological disorders, affecting more than 10 million individuals worldwide and ranking as the second most prevalent neurological condition^1,2^. Genetic factors are discussed to play a pivotal role, suggesting an important genetic etiology of PD^3^. Accordingly, approximately up to 15% of PD cases are associated with causative pathogenic variants in *SNCA*, *LRRK2*, *VPS35*, *RAB32*, *CHCHD2*, *PRKN*, *PINK1*, or *PARK7* or in *GBA1*, the strongest known risk factor for the development of PD^3–7^.

To date, diagnosing PD remains challenging, as existing diagnostic methods primarily assess the extent of motor function decline^8^, which manifests through clinical signs like bradykinesia, resting tremor, rigidity, and postural instability. Therefore, PD is typically diagnosed after its manifestation when up to 80% of the dopaminergic neurons have already been depleted^9,10^. Accurate diagnostic tests are essential, especially in its early, prodromal phase. Molecular markers hold promise, as they may reveal pathological changes even before any signs or symptoms occur. Thus, research has focused on identifying such biomarkers in body fluids like blood, saliva, cerebrospinal fluid (CSF)^11–14^. These efforts have even resulted in the first biological classifications of PD^15,16^. Further, alpha-synuclein seed amplification assays (aSyn-SAAs) have been introduced to improve the diagnostic accuracy of PD^14,17,18^. However, their dependence on invasive CSF sampling limits their use for large-scale screening, emphasizing the need for additional non-invasive approaches.

Notably, there has been a trend towards using several omics approaches like transcriptomics, proteomics and metabolomics studies for identifying biomarkers^19–21^. However, no specific biochemical biomarker has yet been established as definitive diagnostic tool in clinical practice^13,22^. Therefore, it is crucial to propose alternative approaches using sensitive, robust, and clinically applicable analytical techniques. Recent investigations have reported that individuals with PD emit a distinct musky odor, that can be identified in the prodromal phase by hyperosmic individuals^23^. Volatile metabolites in sebum, produced by the skin’s sebaceous glands, may contribute to this signature. Another study found metabolic changes in sebum samples, suggesting a lipid dysregulation in PD^24^. While this discovery appears encouraging, further research is needed to extensively explore the biochemical alterations in this excretory body fluid in PD patients.

Another non-invasively collected yet underexplored excretory biomaterial is breath. Emerging reports suggest that the pathological mechanisms of diseases can significantly influence the composition of breath^25^. Breath analysis has shown promise for diagnosing and monitoring various conditions, including Alzheimer’s disease and PD. However, its application in PD research remains in its infancy. Most studies have predominantly focused on volatile organic compounds (VOCs) using complex (sensor) systems^26–29^, while non-volatile organic compounds (nVOCs) in breath have been largely neglected. Investigating nVOCs could reveal additional biomarkers, offering a comprehensive understanding of breath and disease-related changes. To fill this gap, we recently demonstrated the potential of analyzing the non-volatile metabolome of ‘healthy’ breath using a simple filter-based device^30^.

In this study, we applied this untargeted metabolomic approach using extreme-resolution Fourier-transform ion cyclotron resonance mass spectrometry (FT-ICR-MS) combined with diverse biostatistical analyses (1) to characterize the non-volatile breath-signature of PD patients with and without pathogenic genetic variants in PD-linked genes (2) to provide a comprehensive overview of the altered metabolomic patterns in the breath of diseased versus healthy individuals and (3) to examine potential metabolomic changes in healthy individuals carrying pathogenic variants. This work establishes the first non-volatile metabolomic breath profile in pathogenic or strong coding risk variant (*LRRK2*, *GBA1*, *PRKN*) PD and idiopathic PD (IPD) patients, presenting significant metabolites that may be indicative of PD.

## RESULTS

The exhaled breath sample group consisted of 73 PD patients (female: 23, male: 50, age range 29-84 years) and 90 healthy participants (female: 37, male: 53, age range 20-60 years) with a BMI of all participants ranging from 19.0–25.0 kg m^−2^. Additionally, to validate the properties of breath analysis, metabolic patterns of exhaled breath were compared to those of blood plasma. For this, a blood plasma sample group was recruited, consisting of 91 PD patients (female: 46, male: 45, age range: 29-84) and an independent group of 78 healthy participants (female: 39, male: 39, age range: 25-89) with all participants’ BMI ranging from 19.0-25.0 kg m^−2^. Both patient groups were further categorized into four PD subgroups, including idiopathic PD (no detected pathogenic variant in a known PD gene) and monogenic PD with pathogenic variants in *GBA1*, *LRRK2,* or *PRKN*. Additionally, 4 and 11 non-manifesting carriers were included in the exhaled breath and blood analysis groups, respectively (Table 1). This investigation analyzed 498 exhaled breath and 180 blood plasma samples, comprising 167 and 180 participants, respectively.

**Table 1:** Characteristics of the participants. Participant groups include individuals diagnosed with Parkinson’s disease (PD+), either with a pathogenic or strong coding risk variant in *LRRK2*, *GBA1*, or *PRKN* or without pathogenic variants (idiopathic PD+). Non-manifesting carriers are healthy unaffected individuals carrying a pathogenic variant in one of these genes (PD–), while healthy controls (HC) are unaffected individuals without any known pathogenic variant.

A: Exhaled Breath Analysis
| Characteristic | PD<sup>+</sup> |  |  |  | PD<sup>-</sup> | Healthy controls |
| --- | --- | --- | --- | --- | --- | --- |
|  | *LRRK2* | *GBA1* | *PRKN* | *Idiopathic* | *LRRK2* | No pathogenic variant |
| Overall n=167 | 12 | 35 | 6 | 20 | 4 | 90 |
| Female | 3 | 10 | 4 | 6 | 1 | 37 |
| Male | 9 | 25 | 2 | 14 | 3 | 53 |
| Age [years] | 29-84 (mean: 60±10) |  |  |  | 27-44 (mean: 35±8) | 20-60 (mean: 30±7) |

B: Blood Plasma Analysis
| Characteristic | PD<sup>+</sup> |  |  |  | PD<sup>-</sup> |  | Healthy controls |
| --- | --- | --- | --- | --- | --- | --- | --- |
|  | *LRRK2* | *GBA1* | *PRKN* | *Idiopathic* | *LRRK2* | *GBA1* | No pathogenic variant |
| Overall n=180 | 13 | 35 | 4 | 39 | 8 | 3 | 78 |
| Female | 3 | 11 | 2 | 17 | 3 | 1 | 39 |
| Male | 10 | 24 | 2 | 22 | 5 | 2 | 39 |
| Age [years] | 35-85 (mean: 61±9) |  |  |  | 29-65 (mean: 50±14) |  | 25-89 (mean: 69±15) |

### Core metabolome

In total, 2199 and 3502 chemical formulas and their corresponding putative metabolites were found in exhaled breath and blood plasma samples of all participants. The full sets of annotated metabolites were used to characterize the metabolomic profiles in both body fluids of PD patients. We utilized the Human Metabolome Database (HMDB) ^31^ 2023, which provided corresponding HMDB IDs for metabolites. Before analysis, all metabolites present only in healthy participants were removed from the dataset. As a result, 2898 metabolites detected in blood plasma and 1954 metabolites identified in exhaled breath samples were selected for the metabolite set enrichment analysis (MSEA). Since not every HMDB ID is included in MetaboAnalyst 6.0^32^, the MSEA yielded 1087 and 1475 metabolites for exhaled breath and blood plasma, respectively. These metabolites were categorized into chemical sub-classes (Figure 1). The fraction of the respective chemical class was calculated by dividing the number of detected metabolites by the number of the maximum potential hits. The exhaled breath of PD patients revealed a spectrum of metabolite subclasses originating from diverse metabolic pathways akin to those detected in blood plasma. However, both datasets also display discernible differences regarding metabolite subclasses such as ceramides, fatty amides, fatty acid esters, amino acids, or carbohydrates, which were identified in exhaled breath at relatively lower levels compared to blood plasma. Figure 1 depicts an overview of the metabolomic profile of PD patients and compares the chemical sub-classes found in both body fluids.

**Figure 1:**
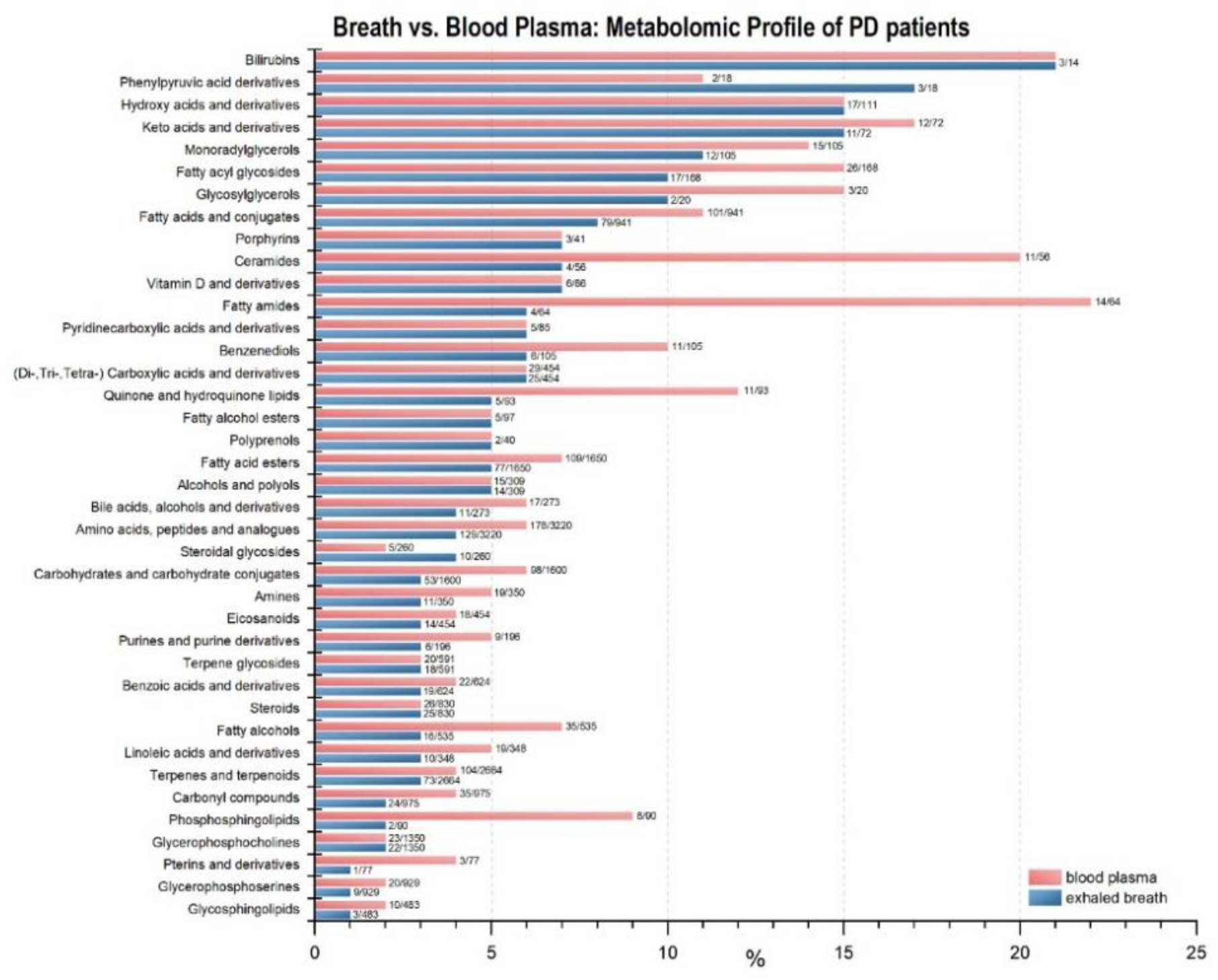
The non-volatile core metabolome of PD patients classified by chemical sub-classes. Core Metabolome analyses were performed using MetaboAnalyst 6.0, employing metabolite set enrichment analysis with 1250 sets of chemical sub-class metabolites. All metabolites present solely in samples of healthy participants were removed from the dataset. Out of 2199 detected metabolites, 1954 were utilized for the metabolome analysis of exhaled breath. Additionally, among the 3502 metabolites identified in blood plasma samples, 2898 metabolites were selected for the analysis. Non-naturally occurring metabolites that are according to Human Metabolome Database (HMDB) part of the human exposome were not excluded. The first value displayed at the bars represents the actual number of detected metabolites in both body fluids while the second value denotes the maximum number of potential metabolites registered in the HMDB. The x-axis summarizes the resulting fraction of the respective chemical class found in both body fluids in percent (%). Only classes accounting for > 1% of the total hits are depicted, ensuring a substantial presence of the classes in the dataset.

### Metabolomic differentiation of PD patients and healthy controls

The datasets were scrutinized to identify PD-specific metabolites distinguishing PD patients, with or without pathogenic variants in known PD genes, from healthy controls. The first analysis compared metabolomic patterns of PD patients and healthy controls (HC) in exhaled breath and blood plasma using volcano analysis, Partial Least Squares Discriminant Analysis (PLS-DA) and random forest classification. This approach revealed discernible differences in the metabolomic profiles of PD patients and healthy controls.

Figure 2 summarizes the results obtained from all three statistical approaches. The volcano analysis of breath samples revealed 89 significant hits differing between PD patients and HC (FC > 2.0, adjusted *p*-value (FDR correction) < 0.1) (Figure 2A). Additionally, scores plot from PLS-DA analysis exhibited a clear separation between PD and HC clusters (R2Y = 0.991 and Q2Y = 0.961 for five components) (Figure 2B). Lastly, employing random forest classification, the algorithm categorized the dataset with notable accuracy (Figure 2C). All HC were correctly assigned, while 72 of 73 PD patients were categorized as such with a measured out-of-bag (OOB) error rate of < 1%.

**Figure 2:**
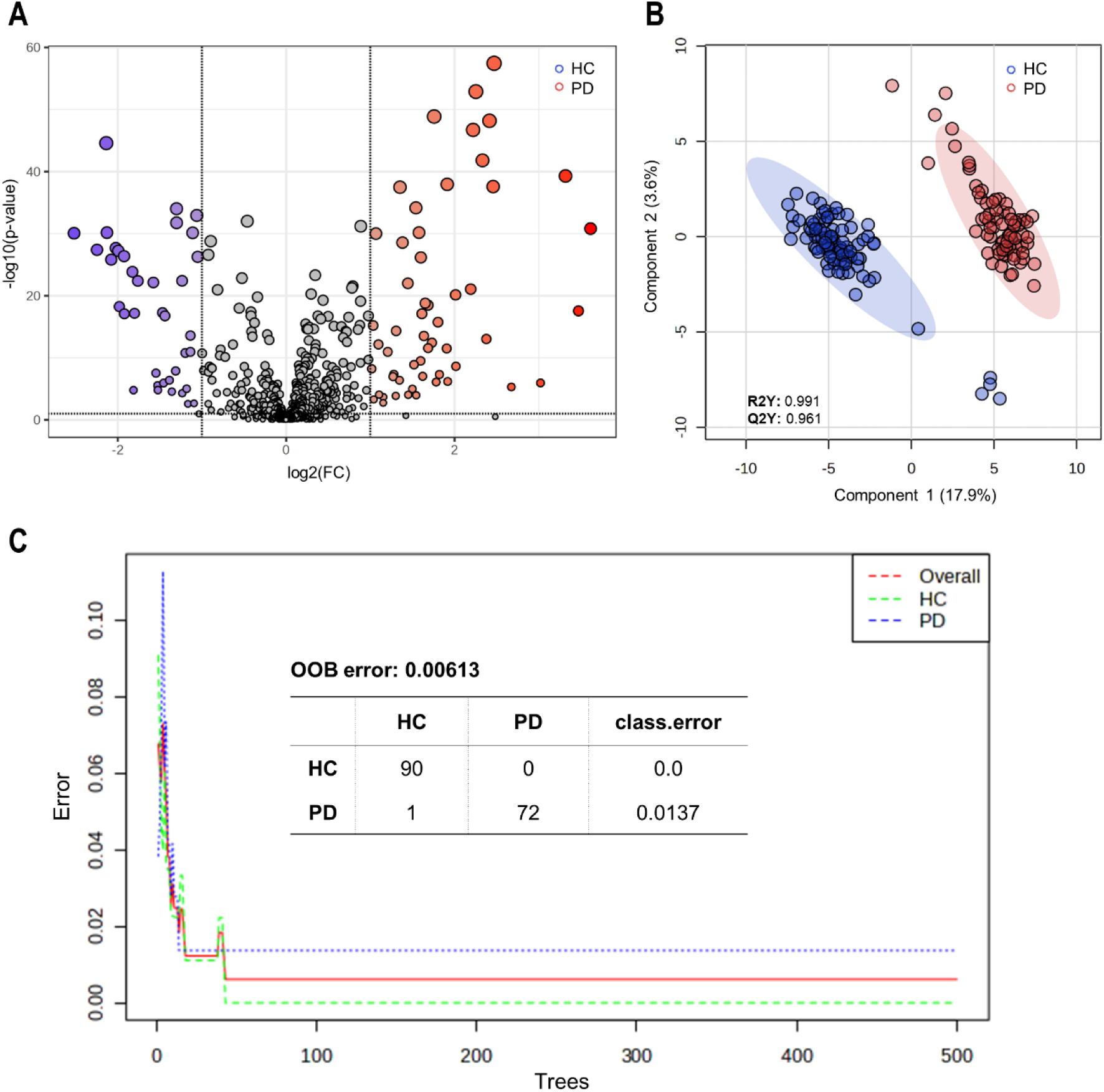
Elaboration of metabolic patterns in exhaled breath using three biostatistical methods. **A** Univariate analysis: 89 metabolites identified by volcano plot differing between PD patients and healthy controls (HC), meeting the criteria of a fold change (FC) threshold > 2.0 and an adjusted *p*-value (FDR correction) of < 0.1. Metabolites exhibiting a log2(FC)> 0 (red) indicate elevated intensities in PD patients, while those with a log2(FC) < 0 (blue) signify higher levels in HC. **B** Multivariate analysis: Scores plot of PLS-DA analysis depicting a clear separation of the metabolite profile of PD patients (red) and HC (blue) with R2Y= 0.991 and Q2Y= 0.961 for five components. **C** Classification analysis: Random forest classification with 40 predictors and 500 trees. Based on their metabolite profile, 72 of 73 PD patients as well as all HC were correctly classified, resulting in an out-of-bag (OOB) error of 0.00613. All calculations and analyses were carried out using MetaboAnalyst 6.0.

Employing the same methodology and biostatistical analyses for blood plasma samples yielded comparable results. All three methods distinguished the metabolomic profiles of PD patients from HC (Supplementary Figure 1). Supplementary Figure 1A shows the volcano analysis of blood plasma samples, identifying 82 significantly different metabolites between PD patients and HC (FC > 2.0, adjusted *p*-value (FDR correction) < 0.1). Similar to the exhaled breath analysis, the PLS-DA scores plot generated demonstrated a distinct cluster separation between PD and HC (R2Y = 0.984 and Q2Y = 0.921 for five components) (Supplementary Figure 1B). The random forest algorithm correctly classified all HC and PD patients based on their metabolite profiles, achieving a classification rate of 100%. To summarize, all three tests showed discernible differences in metabolic patterns between PD patients and healthy individuals in both body fluids.

Furthermore, to cross-validate the biostatistical methods and to pinpoint key metabolites contributing to classification of PD patients and HC, we compared the top 15 metabolites from PLS-DA (VIP Score > 2.5) and random forest analysis (mean decrease accuracy > 0.001) (Supplementary Figure 2 and 3) in both fluids. None of the top 15 exhaled breath metabolites were detected in blood plasma using PLS-DA (Supplementary Figure 3A) and Random Forest (Supplementary Figure 3B).

To further evaluate the data-driven PD prediction, we conducted an additional multivariate exploratory ROC curve analysis for both exhaled breath (Figure 3) and blood plasma (Supplementary Figure 4). Figure 3A and Supplementary Figure 4A illustrate the ROC curve analyses for independent metabolite features (*n* = 5, 10, 15, 25, 50 and 100). Comparing the ROC curves, both models classified PD patients and HCs with AUCs of 0.99-1 (CI 95%). Accordingly, both datasets predicted PD accurately (> 96%) when considering only 10 metabolite features (Figure 3B, Supplementary Figure 4B). While adding more features showed enhanced prediction accuracy, the results indicate that up to 25 features were already sufficient for a reliable prediction of PD in both body fluids. In breath analysis, all 73 PD patients were accurately predicted solely based on the metabolomic pattern, utilizing only 10 features (Figure 3C). Similarly, when evaluating blood plasma, all HCs were correctly predicted, while only 5 out of 91 PD patients were misclassified. This result was achieved using 25 features selected as the best classifier model according to the AUC (Supplementary Figure 4C). Comparing these results with those obtained from random forest classification (Figure 2C, Supplementary Figure 1C), both statistical approaches accurately distinguished PD patients from HCs.

**Figure 3:**
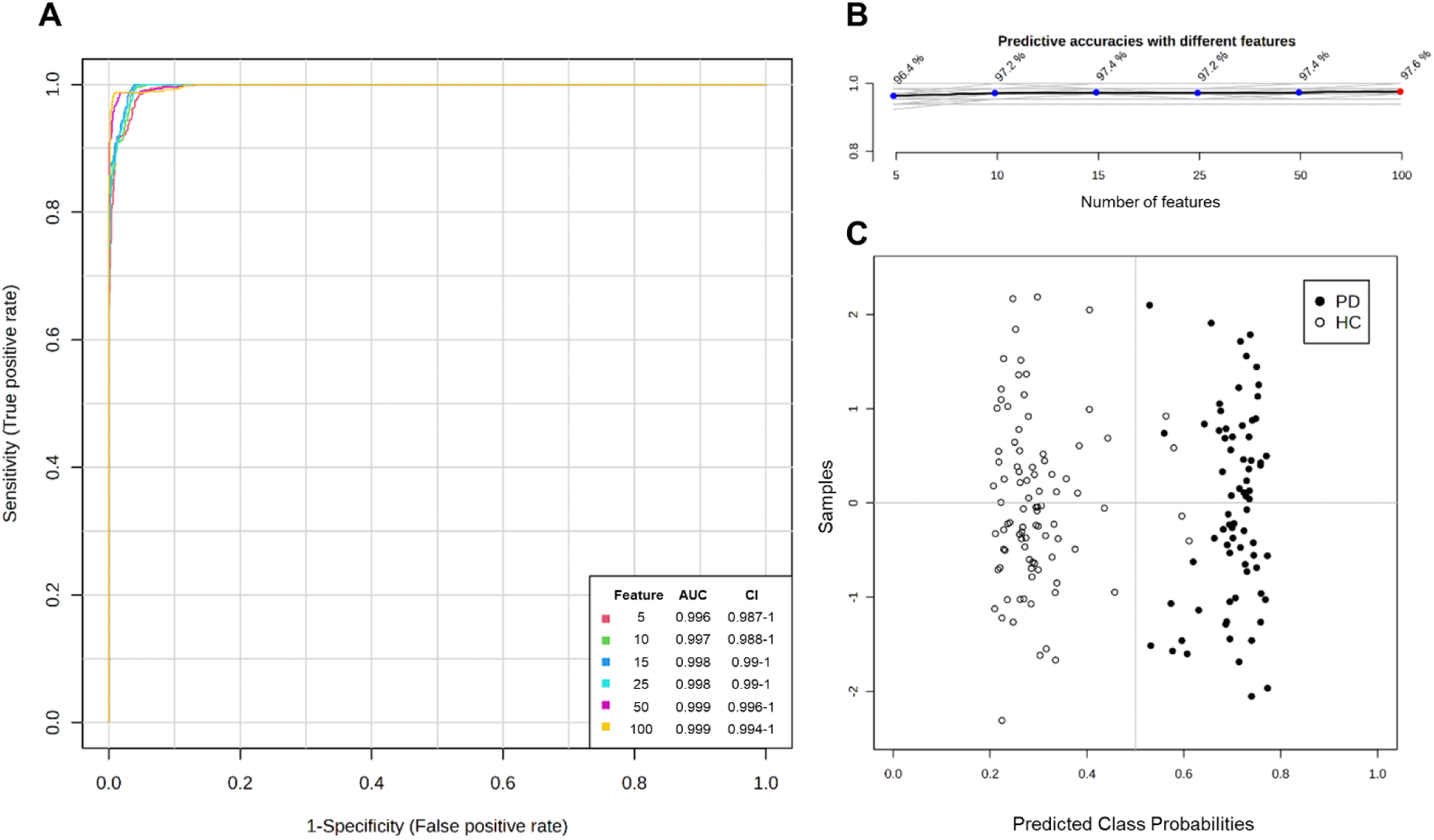
Data-based PD prediction via multivariate exploratory ROC curve analysis of exhaled breath samples. PLS-DA and its built-in feature was selected as the classification method and the ranking method, respectively. The analysis was conducted using two latent variables. **A** ROC curve analyses for *n* = 5, 10, 15, 25, 50 and 100 independent metabolite features (without combining) in PD patients and healthy controls (HC). AUC and 95% confidence intervals (CI) were calculated by Monte Carlo cross validation (MCCV) using balanced sub-sampling **B** Predictive accuracies (y-axis) in percent (%) for *n* = 5, 10, 15, 25, 50 and 100 independent metabolite features. **C** Predicted class probabilities using the best classifier (n=10) based on the AUC. Using this model, all PD patients (n=73) were accurately predicted while four of 90 healthy individuals were classified as PD patients, (currently) false positively. Same results were obtained with n ≥ 15 features. Since a balanced sub-sampling technique was implemented during model training, the classification boundary consistently aligns with x = 0.5, indicated by the dotted line. ROC curve analyses were carried out using MetaboAnalyst Biomarker Analysis 6.0.

After validating the results obtained by biostatistical methods, the significant metabolites responsible for this differentiation were examined. To explore features identified by multiple statistical methods across both body fluids, we conducted a Venn analysis, combining all significant metabolites found in volcano analysis, PLS-DA and random forest analysis (Figure 4). This investigation yielded 89 metabolites in exhaled breath and 82 in blood plasma different between PD and HC identified across all three statistical tests (Figure 4A). No metabolites were exclusive to any single test. Rather, 500 and 837 metabolites were commonly identified between the PLS-DA and random forest analyses for exhaled breath and blood plasma, respectively.

**Figure 4:**
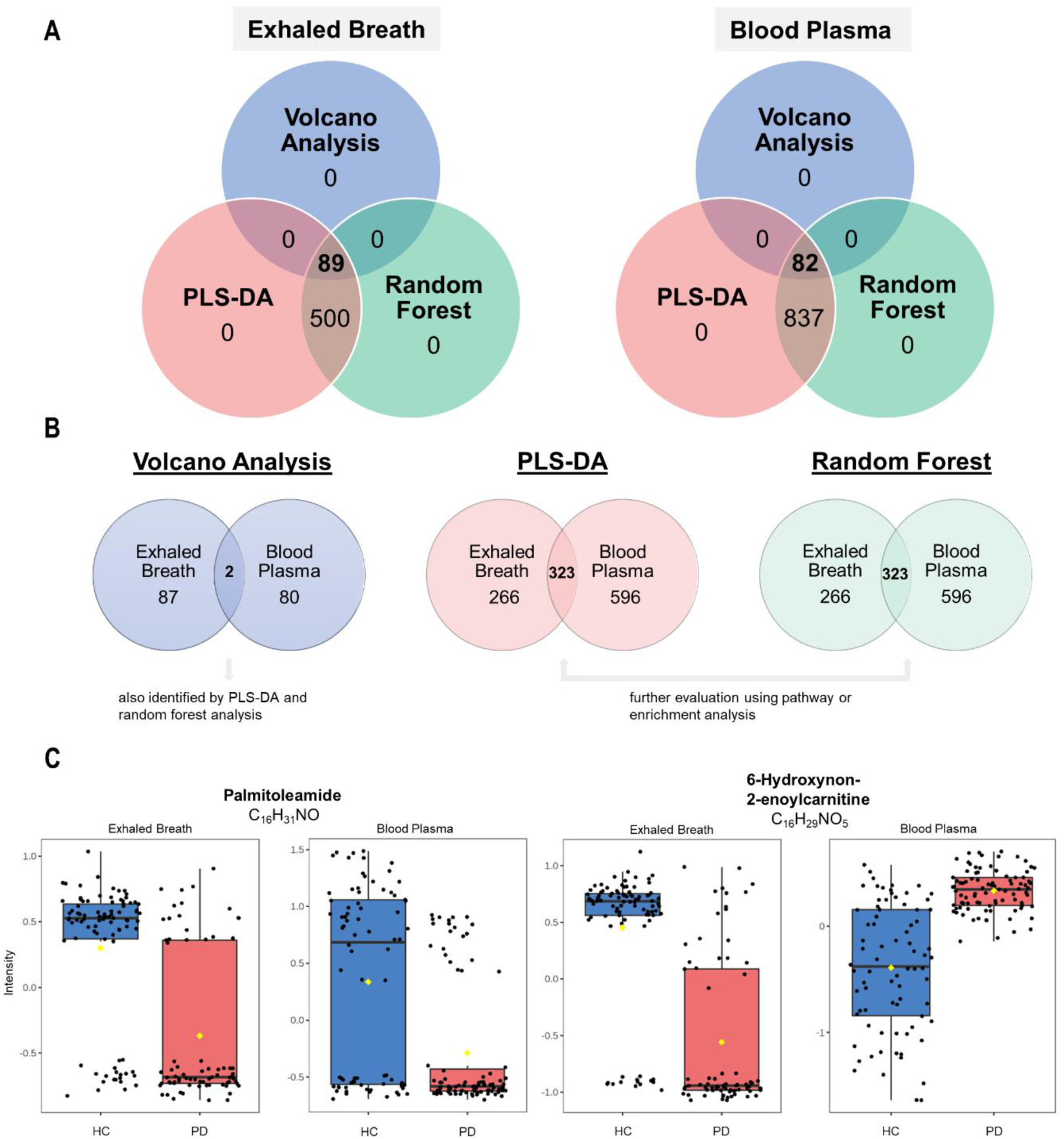
Venn analysis and comparison of significant metabolites identified in all tests. **A.** The significant results obtained from all three tests (volcano analysis, PLS-DA, random forest classification) combined in a Venn diagram resulted in 89 and 82 significant metabolites, commonly identified in all three tests in exhaled breath and blood plasma, respectively. **B** A second Venn analysis comparing those aforementioned hit features between exhaled breath and blood plasma reveals two common metabolites significantly different between PD patients and healthy controls (HC) in all tests. 323 common metabolites were detected exclusively in two of the three tests (PLS-DA, random forest) in both body fluids. **C** Box plots of the two significant metabolites with chemical formula and putative identity found across all biostatistical approaches in both, exhaled breath (left) and blood plasma (right), respectively. The results were significant between PD patients (red, n=73 biologically independent exhaled breath and n=91 biologically independent plasma samples) and healthy controls (HC) (blue, n=90 biologically independent exhaled breath and n=78 biologically independent plasma samples). Black dots represent the values from all samples. The box and whiskers summarize the normalized values, with the center line presenting the median. The mean value is indicated as a yellow diamond.

To explore potential interfluidal overlaps, a subsequent Venn analysis was conducted. This analysis compared the results from the three statistical tests for both fluids (Figure 4B) to identify shared biomarkers between breath and blood plasma that could contribute to a unified PD-specific metabolomic profile. This examination consistently identified 323 significant metabolites using PLS-DA and random forest for both body fluids. However, a substantial number of the metabolites detected in body fluids is unique to a particular matrix, with 266 exclusive to exhaled breath and 596 to blood plasma. Only palmitoleamide (C_16_H_31_NO) and 6-hydroxynon-2-enoylcarnitine (C_16_H_29_NO_5_) were commonly detected across all statistical tests in both exhaled breath and blood plasma (Figure 4C). However, these shared metabolites varied in distribution between body fluids. While palmitoleamide (C_16_H_31_NO) exhibited consistent results, 6-hydroxynon-2-enoylcarnitine (C_16_H_29_NO_5_) showed higher intensity levels in PD blood plasma but lower in exhaled breath samples when comparing patients to controls. From the Venn analysis, we selected the 15 most robust metabolites (p value << 0.001, VIP scores > 1.5, mean decrease accuracy > 0.0001) and calculated the expression ratios between both groups. Table 2 and 3 outline these metabolites for exhaled breath and blood plasma, respectively. The top 15 metabolites differed between the two body fluids. Seven metabolites (docosanamide, nonadecanoic acid, tricosanoic acid, sorbitan palmitate, 2-hydroxy-3-methyl-pentanoic acid, 2-methyl-3-hydroxy-butyric acid, and ubiquinone-1) (Table 2) were among the 323 commonly detected in both exhaled breath and blood plasma (Figure 4B) but were not among the top significant hits in blood plasma. Similarly, only 3 of the top 15 blood plasma metabolites (C_9_H_7_NO, C_14_H_22_N_2_O and 3-methoxytyrosine) (Supplementary Table 1), were detected among the 323 shared metabolites.

**Table 2:** The 15 most relevant metabolites in exhaled breath of PD patients obtained from Venn analysis. The metabolites were identified in all three biostatistical methods with expression ratios obtained from a fold change analysis (FC=2.0), adjusted *p*-values << 0.001 (volcano analysis), VIP scores >1.5 (PLS-DA) and mean decrease accuracy values >0.0001 (random forest), with arrows indicating either higher or lower levels in PD patients compared to healthy controls. Metabolites labelled with a hash mark (#) were identified in Venn analysis of blood plasma as well. MG: monogylceride.

| Chemical formula | Putative identity | Expression ratio (FC) | Volcano plot (adj. *p*-value) | PLS-DA (VIP score) | Random forest (MDA) |
| --- | --- | --- | --- | --- | --- |
| C<sub>22</sub>H<sub>45</sub>NO | Docosanamide# | ↑ 0.18 | 4.45E-62 | 3.16 | 0.015 |
| C<sub>20</sub>H<sub>40</sub>O<sub>2</sub> | Eicosanoic acid | ↑ 0.21 | 5.46E-57 | 2.98 | 0.023 |
| C<sub>19</sub>H<sub>38</sub>O<sub>2</sub> | Nonadecanoic acid# | ↑ 0.30 | 7.59E-53 | 2.56 | 0.024 |
| C<sub>21</sub>H<sub>42</sub>O<sub>4</sub> | Octadecanoyl MG | ↑ 0.19 | 5.49E-52 | 3.22 | 0.025 |
| C<sub>18</sub>H<sub>36</sub>O<sub>2</sub> | Stearic acid | ↑ 0.22 | 3.13E-50 | 2.99 | 0.020 |
| C<sub>23</sub>H<sub>46</sub>O<sub>2</sub> | Tricosanoic acid# | ↑ 0.20 | 3.77E-45 | 3.14 | 0.029 |
| C<sub>22</sub>H<sub>42</sub>O<sub>6</sub> | Sorbitan palmitate# | ↑ 0.10 | 4.49E-42 | 3.92 | 0.011 |
| C<sub>22</sub>H<sub>44</sub>O<sub>4</sub> | Nonadecanoyl MG | ↑ 0.27 | 3.14E-41 | 2.68 | 0.028 |
| C<sub>16</sub>H<sub>32</sub>O<sub>2</sub> | Palmitic acid | ↑ 0.40 | 5.37E-41 | 2.15 | 0.007 |
| C<sub>24</sub>H<sub>48</sub>O<sub>2</sub> | Tetracosanoic acid | ↑ 0.18 | 1.41E-40 | 3.23 | 0.029 |
| C<sub>19</sub>H<sub>38</sub>O<sub>4</sub> | Hexadecanoyl MG | ↑ 0.35 | 1.10E-37 | 2.32 | 0.006 |
| C<sub>6</sub>H<sub>12</sub>O<sub>3</sub> | 2-Hydroxy-3-methyl-pentanoic acid# | ↓ 2.48 | 6.19E-37 | 2.20 | 0.031 |
| C<sub>5</sub>H<sub>10</sub>O<sub>3</sub> | 2-Methyl-3-hydroxy-butyric acid# | ↓ 2.48 | 1.46E-34 | 2.07 | 0.016 |
| C<sub>17</sub>H<sub>34</sub>O<sub>2</sub> | Heptadecanoic acid | ↑ 0.48 | 4.08E-33 | 1.84 | 0.003 |
| C<sub>14</sub>H<sub>18</sub>O<sub>4</sub> | Ubiquinone-1# | ↓ 4.43 | 3.75E-32 | 2.70 | 0.002 |

This investigation aimed to identify not only the most statistically robust but also the most relevant metabolites as potential biomarker candidates. Therefore, the analysis proceeded with a univariate classical ROC analysis of the top 15 metabolites with an AUC > 0.96. Figure 5 exhibits boxplots of the top 10 significant metabolites identified through all statistical analyses in exhaled breath. Seven of the top 10 metabolites, including tetracosanoic acid, tricosanoic acid, eicosanoic acid, nonadecanoic acid, homophytanic acid, stearic acid, palmitic acid, exhibit elevated levels in PD patients. To ascertain the significance of these metabolites in PD patients, irrespective of pathogenic variants, we performed a one-way ANOVA with Tukey HSD correction (*p*-value < 0.05). The results reveal that all these metabolites show significantly higher levels across all subgroups compared to HC.

**Figure 5:**
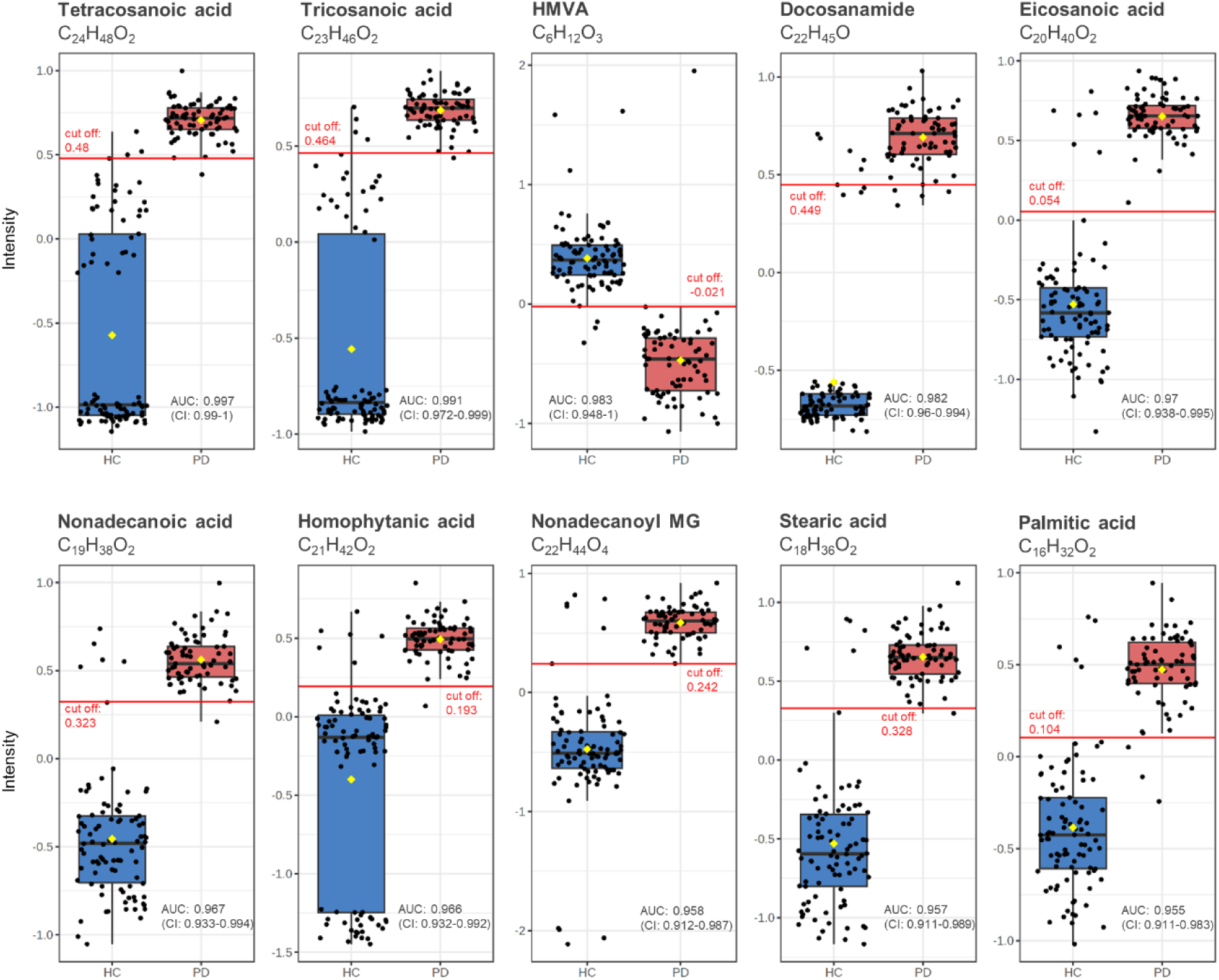
The top 10 significant metabolites identified in exhaled breath using univariate ROC curve analyses. The univariate ROC analysis used 500 bootstrappings for calculating the 95% confidence interval (CI). Only those metabolites were selected that were identified in Venn analysis across volcano analysis (FC > 2.0, adjusted *p*-value (FDR correction) < 0.1), PLS-DA (VIP Score > 2.5) and random forest analysis (mean decrease accuracy > 0.001). The box plots depict significant differences between the group of PD patients (red, n=73 biologically independent samples) and healthy controls (HC) (blue, n= 90 biologically independent samples). Black dots represent the values from all samples. The box and whiskers summarize the normalized values with the center line presenting the median. The mean value is indicated as a yellow diamond. The red line denotes the optimal cut-off values (closest to the top-left corner) obtained from univariate ROC analysis. MG: monoglyceride. HMVA: 2-Hydroxy-3-methylvaleric acid.

None of these 10 metabolites were found among the 82 significant metabolites of blood plasma identified through Venn analysis (Figure 4A). However, tricosanoic acid, HMVA (2-Hydroxy-3-methylvaleric acid), nonadecanoic acid and homophytanic acid were found among the 323 shared metabolites between exhaled breath and blood plasma. Re-evaluating the top 15 metabolites of blood plasma (Supplementary Table 1) with an additional univariate classical ROC analysis, merely 4 of these metabolites (C_12_H_26_O, C_2_H_7_O_3_P, C_14_H_22_N_2_O, C_12_H_24_O) exhibited an AUC greater than 0.96.

Performing various statistical analyses revealed significant differences between PD and HC groups. However, the PD group was not stratified by mutation status in these aforementioned analyses. To assess the impact of pathogenic variants on the observed metabolic profile, PD patients were divided into two groups: idiopathic PD (IPD) and PD with mutation (mPD). Statistical tests were conducted for both groups compared to HC. The results were consistent, with identical distributions when comparing the top 20 outcomes from volcano plots, PLS-DA, and random forest analyses, similar to the overall PD and HC comparison.

### Analysis of metabolomic patterns in PD patients with and without genetic variants

Metabolomic patterns in PD patients were examined for patients with and without genetic variants in known PD genes like *GBA1*, *LRRK2* and *PRKN*. Accordingly, we performed statistical analyses to compare IPD patients with those carrying one of the aforementioned variants (mPD) (Supplementary Information). The volcano analysis identified 10 and 23 significant metabolites in the breath and blood plasma samples of the IPD and mPD group (Supplementary Figures 5 and 6). These particular metabolites were not identified as significantly different when comparing all PD patients to HC. When selecting only naturally occurring metabolites, the volcano plot highlighted N-decanoylglycine (C_12_H_23_NO_3_) and 2-octenoylcarnitine (C_15_H_27_NO_4_) as significantly elevated in breath of the mPD group compared to IPD patients (Supplementary Figure 5).

Further multivariate analysis and classification of all PD subgroups were conducted. Supplementary Figure 7 summarizes the PLS-DA and random forest classification outcomes. PLS-DA revealed a clear separation of PD patients with and without pathogenic variants in both body fluids (Supplementary Figure 7A). Additionally, the random forest accurately classified all IPD patients in blood plasma and exhaled breath, with only one exception, while misclassifying 3 of 53 PD mutation carriers in breath and 1 of 51 in blood. Assessing all PD subgroups (IPD, PD_ *GBA1*, PD_*LRRK2*, and PD_*PRKN*), the scores plot exhibited a better cluster separation in blood plasma compared to breath (Supplementary Figure 7B). The random forest analysis effectively classified IPD patients and PD patients carrying a *GBA1* mutation in both fluids but showed lower accuracy for *LRRK2* and *PRKN* subgroups, increasing OOB errors in both exhaled breath (0.16) and blood plasma (0.09) (Supplementary Figure 7B).

### Evaluation of healthy individuals with a pathogenic variant (non-manifesting LRRK2 mutation carriers)

Subsequent investigations focused on the small group of 4 healthy individuals harboring a pathogenic *LRRK2* variant. Initially, a multivariate ROC curve analysis was employed for classification (Figure 6). The exhaled breath analysis exhibited higher ROC AUC values compared to blood plasma, with respective values of 0.98 and 0.94 for 25 features (Figure 6A). This analysis discriminated the 4 unaffected individuals carrying a pathogenic variant in *LRRK2* from other HC. When examining the predicted class probabilities, 5 of 90 healthy individuals were misclassified as mutation carriers in exhaled breath, while 10 of 78 were misclassified in the blood plasma group (Figure 6B). However, this model accurately predicted all HC individuals with an *LRRK2* variant (exhaled breath: n=4 and blood plasma: n=8).

**Figure 6:**
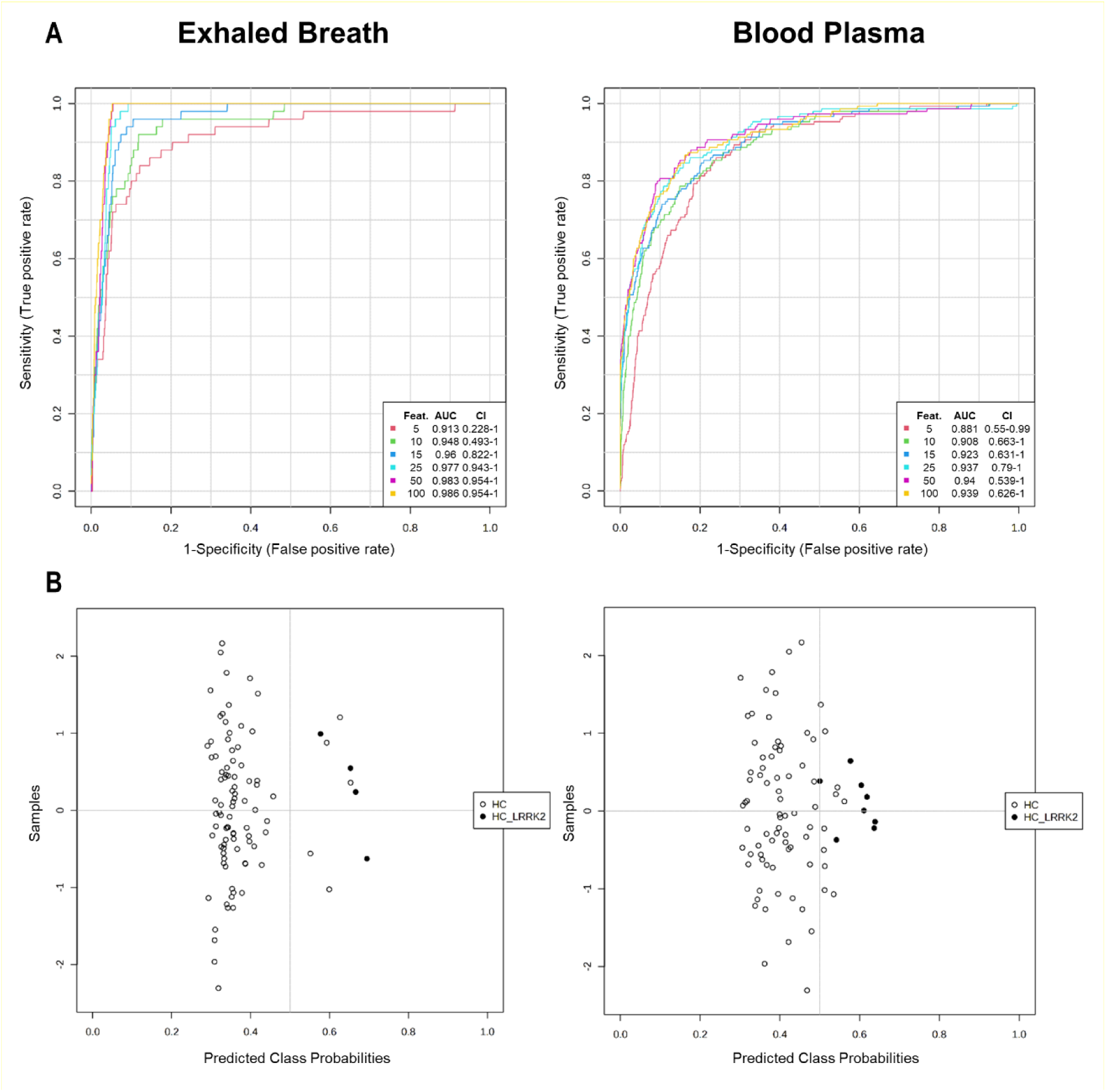
Multivariate exploratory ROC curve analyses of exhaled breath vs. blood plasma for classifying healthy controls with and without *LRRK2* mutation. PLS-DA and its built-in feature was selected for the classification method and the ranking method, respectively. The analysis was conducted using two latent variables. **A** Comparison of ROC curve analyses of exhaled breath and blood plasma for *n* = 5, 10, 15, 25, 50 and 100 independent metabolite features in healthy controls and healthy individuals with *LRRK2* mutation. AUC and 95% confidence intervals (CI) were calculated by Monte Carlo cross validation (MCCV) using balanced sub-sampling **B** Predicted class probabilities using the best classifier (exhaled breath: n=10, blood plasma: n=15) based on the AUC. Using this model, all 4 and 8 healthy controls carrying a *LRRK2* mutation were accurately predicted and differed from healthy controls by metabolite patterns in exhaled breath and blood plasma, respectively. Since a balanced sub-sampling technique was implemented during model training, the classification boundary consistently aligns with x = 0.5, indicated by the dotted line. ROC curve analyses were carried out using MetaboAnalyst Biomarker Analysis 6.0.

Additionally, PLS-DA, volcano plot, random forest and a classical univariate ROC curve analysis were conducted to identify significant metabolites differing between the group of non-manifesting *LRRK2* mutation carriers and HC. Table 3 presents the top metabolites detected in exhaled breath when applying the aforementioned tests (*p*-value < 0.001, VIP Score > 1.5, mean decrease accuracy > 0.0001 and ROC AUC > 0.98).

**Table 3:** Top metabolites in breath of unaffected participants carrying a *LRRK2* mutation. Metabolites were identified across various biostatistical tests (PLS-DA, volcano plot, random forest), as well as in biomarker analysis using univariate ROC curve analyses. Only metabolites meeting the criteria of a VIP Score > 1.5, *p*-value < 0.001, mean decrease accuracy > 0.0001, and a ROC AUC > 0.98 were selected. The metabolites were further analyzed based on their expression ratios obtained from a fold change analysis (FC=2). Arrows indicate either higher or lower levels in healthy participants with *LRRK2* mutation compared to healthy participants without mutation. Metabolites labelled with an asterisk (*) were also identified and characterized among the top 10 metabolites differing between PD patients and healthy controls with a similar distribution. MG: monogylceride.

| Chemical formula | Putative identity | Expression ratio (FC) |
| --- | --- | --- |
| C<sub>21</sub>H<sub>42</sub>O<sub>4</sub> | **Octadecanoyl MG** | ↑ 0.11 |
| C<sub>18</sub>H<sub>36</sub>O<sub>2</sub> | Stearic acid* | ↑ 0.15 |
| C<sub>20</sub>H<sub>40</sub>O<sub>2</sub> | Eicosanoic acid* | ↑ 0.19 |
| C<sub>19</sub>H<sub>38</sub>O<sub>4</sub> | **Hexadecanoyl MG** | ↑ 0.19 |
| C<sub>19</sub>H<sub>38</sub>O<sub>2</sub> | Nonadecanoic acid* | ↑ 0.23 |
| C<sub>22</sub>H<sub>44</sub>O<sub>4</sub> | Nonadecanoyl MG* | ↑ 0.19 |
| C<sub>26</sub>H<sub>38</sub>O<sub>8</sub> | **1-Methylene-5alpha-androstan-3alpha-ol-17-one glucuronide** | ↑ 0.18 |
| C<sub>16</sub>H<sub>32</sub>O<sub>2</sub> | Palmitic acid* | ↑ 0.27 |
| C<sub>21</sub>H<sub>42</sub>O<sub>2</sub> | Homophytanic acid* | ↑ 0.32 |
| C<sub>17</sub>H<sub>34</sub>O<sub>2</sub> | **Heptadecanoic acid** | ↑ 0.29 |
| C<sub>25</sub>H<sub>39</sub>O<sub>11</sub>P | **PA(2:0/20:5(7Z,9Z,11E,13E,17Z)-3OH(5,6,15))** | ↑ 0.39 |
| C<sub>6</sub>H<sub>12</sub>O<sub>3</sub> | 2-Hydroxy-3-methylvaleric acid (HMVA)* | ↓ 2.56 |
| C<sub>5</sub>H<sub>10</sub>O<sub>3</sub> | **(2S,3R)-3-Hydroxy-2-methylbutanoic acid** | ↓ 2.17 |

The metabolites presented in Table 3 were compared to those found in the comparison between PD and HC. Notably, 7 out of the top 10 metabolites identified as different between PD and HC (Figure 5) were also significantly different in non-manifesting *LRRK2* carriers compared to HC.

Moreover, non-manifesting *LRRK2* carriers exhibited elevated monoglycerides (MG) levels (Table 3). A one-way ANOVA of MG levels across PD subgroups revealed that *LRRK2*-PD patients showed significantly higher levels compared to other PD subgroups (Figure 7). MG levels were also found significantly higher in PD patients compared to HC, irrespective of the pathogenic variants.

**Figure 7:**
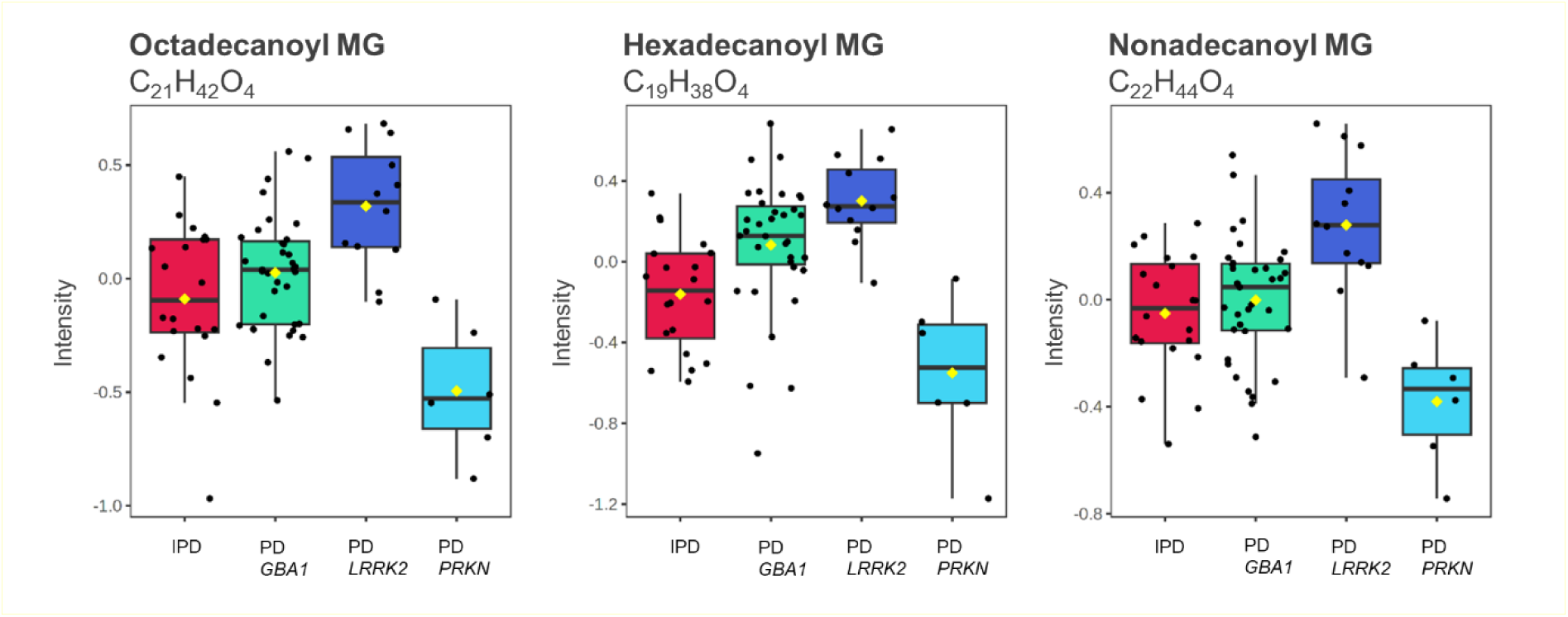
Box plots of three significant metabolites identified in exhaled breath of PD subgroups. Significant findings obtained from a one-way ANOVA combined with a Tukey honestly significant difference (HSD) test for FDR correction (*p*-value <0.05). Differences were observed between all PD subgroups (1) IPD patients (red, n=20 biologically independent exhaled breath samples), (2) PD_ *GBA1* (green, n= 35 biologically independent exhaled breath samples), (3) PD_*LRRK2* (blue, n=12 biologically independent exhaled breath samples) and (4) PD_*PRKN* (turquoise, n=6 biologically independent exhaled breath samples). The PD patients with a *LRRK2* mutation showed elevated levels compared to the other PD subgroups. All three metabolites showed elevated levels in PD patients compared to healthy controls. Black dots represent the values from all samples. The box and whiskers summarize the normalized values with the center line presenting the median. The mean value is indicated as a yellow diamond. These three metabolites were also found significant in a univariate ROC curve analysis comparing healthy controls to healthy individuals carrying a *LRRK2* mutation (see Table 3). MG: monoglyceride.

The same analysis using blood plasma from non-manifesting *LRRK2* carriers revealed that none of the top 20 blood plasma metabolites matched the significant differences found between PD patients and HC (Supplementary Table 1). However, the volcano analysis identified 117 metabolites differing between non-manifesting carriers and HC (p < 0.001), while PLS-DA and random forest classification detected 317 and 242 metabolites, respectively (VIP Score > 1.5, mean decrease accuracy > 0.0001).

Among the top 20 differentiating metabolites between PD and HC, only octadecanamide (C_18_H_37_NO) stood out in the top hits of a univariate ROC curve analysis (ROC AUC 0.94) comparing non-manifesting *LRRK2* carriers and HC. It showed higher levels in HC with a fold change of 2.45. However, this metabolite did not exhibit significance specifically for PD patients with an *LRRK2* mutation (one-way ANOVA), yet it demonstrated significantly higher levels in the HC group compared to all PD patients.

## DISCUSSION

This breathomics study applied a non-targeted metabolomic approach focusing on the non-volatile organic compounds in breath, as previously used for healthy individuals^30^. Utilizing ultra-high-resolution FT-ICR-MS facilitated an extremely high sensitivity for distinguishing traces within femtomole range. Here, we depict the non-volatile metabolomic profile, a breathome, of PD patients, both with and without pathogenic variants in known PD genes, presenting metabolite patterns differing from those of healthy controls. This discussion mainly focuses on exhaled breath analysis while using the commonly employed blood plasma as a reference to compare the body fluids.

The pre-processed datasets successfully yielded 2199 and 3502 chemical formulas and their corresponding putative metabolites in exhaled breath and blood plasma of all participants, respectively. When assigning identities to metabolites, it is important to consider the large number of isobaric compounds sharing the same chemical formula. If multiple putative identities were assigned, we selected the first listed metabolite of the annotation list. However, for all proposed significant metabolites, plausible alternatives within the annotation list were also explored. Such feasible alternatives will be discussed later in the discussion. In general, blood plasma yielded over 1000 more metabolites compared to breath. However, it should be considered that the 2199 metabolites detected in breath selectively represent only its non-volatile fraction. Hence, a breathomics approach analyzing both non-volatile and volatile organic compounds might identify even more metabolites. Applying a metabolite set enrichment analysis (MSEA) enabled the characterization of the core metabolome, identifying 1087 metabolites for exhaled breath and 1475 for blood plasma. These metabolites were successfully categorized into chemical sub-classes (Figure 1), highlighting biochemically versatile molecules with various functions in the human body. Blood plasma appears to be richer in certain chemical sub-classes, revealing a greater number of metabolites from specific sub-classes such as but not limited to ceramides, fatty amides, fatty acid esters, amino acids, and peptides, as well as carbohydrates. Notably, the number of carbohydrates and ceramides is nearly double in blood plasma compared to exhaled breath. Despite those differences, other sub-classes such as fatty alcohol esters, steroids, glycerophosphocholines, hydroxyl acids, and vitamin D and derivatives evidently exhibit similar proportions in both body fluids. Comparing exhaled breath and blood plasma analyses shows that breath is just as suitable for metabolomics approaches as blood plasma. The striking resemblance in their core metabolome composition further reveals that breath analysis provides valuable insights without loss of information. Remarkably, non-invasive breath sampling enables the identification of similar chemical classes from just 30 exhalations. The presented chemical sub-class groups in Figure 1 encompass all metabolites found in two body fluids of PD patients, with the presented metabolites not necessarily depicting the significant differences compared to healthy controls. These differences become more apparent in statistical analyses, which are necessary to identify any distinctive signatures in PD patients.

Remarkably, similar to blood plasma, exhaled breath analysis effectively differentiated between healthy individuals and those with PD based on their metabolomic profile, regardless of the biostatistical test applied (Figure 2, 3 and Supplementary Figure 1, 4). These findings highlight the potential of exhaled breath as a non-invasive alternative biomaterial for metabolomics studies and potential diagnostic applications. The results obtained from the Venn analysis further support this conclusion. While there is a significant overlap of metabolites found in both body fluids (n=323), a considerable portion is unique to each fluid. Specifically, 266 metabolites were detected exclusively in exhaled breath and 596 solely in blood plasma (Figure 4B). This suggests that exhaled breath provides unique insights into metabolomic investigations, offering a diverse set of information not accessible from commonly used body fluids such as blood plasma.

When examining the significant hits shared between both fluids, two hits consistently emerged as significant across all statistical methods, distinguishing between healthy and diseased individuals (Venn analysis, Figure 4B). One of the two metabolites is palmitoleamide (C_16_H_31_NO), an amide of palmitoleic acid (monounsaturated fatty acid, 16:1n-7). This metabolite exhibited significantly lower levels in PD patients in both exhaled breath and blood plasma samples. It was first discovered as one of five endogenous primary fatty acid amides (PFAM) in luteal phase plasma with their physiological significance being unclear^33^. However, reports suggest that PFAM play a significant role as cell signaling lipids in the mammalian nervous system^34,35^. Since the biological active oleamide, the most studied PFAM, has been reported as a natural component of cerebrospinal fluid (CSF)^35^, it is likely that other PFAMs are also present in CSF, each with distinct roles as signaling lipids. It is conceivable that the concentration and composition of PFAMs may change in PD, leading to alterations in CSF compared to that of healthy individuals. Furthermore, the second metabolite commonly found significant belongs to the class of acylcarnitines. 6-hydroxynon-2-enoylcarnitine (C_16_H_29_NO_5_) was found higher elevated in PD patients, however, showed a contrary distribution in exhaled breath (Figure 4C). Interestingly, decreased levels of long-chain acylcarnitines have previously been discussed as potential biomarkers for PD^36^. Assessing the top 15 significant hits identified in the respective fluid via Venn analysis revealed that none of the top metabolites were congruent (Table 2, Supplementary Table 1). Despite a similar core metabolome composition, different significant metabolites from various chemical classes can emerge in both fluids, suggesting notable differences between the two biomaterials.

An in-depth biostatical analysis of exhaled breath identified the top 10 metabolites that differentiate between PD patients and healthy individuals (Figure 5). These metabolites were not found among the top hits in blood plasma. Cross-validated through multiple statistical tests, they appear to be the most robust and relevant markers for distinguishing the metabolomic profiles of the two groups. Interestingly, 9 of 10 are linked directly or indirectly to the lipid metabolism. Docosanamide (C_22_H_45_O) another fatty acid amide, was found higher abundant in PD patients compared to healthy controls. While this metabolite has not been previously linked to PD, however, as a member of the primary fatty acid amide family^37^, it might serve as signaling lipid in the nervous system as well^34,35^. Furthermore, the glycerolipid nonadecanoyl monoglyceride (MG) (C_22_H_44_O_4_) was elevated in PD patients as well. The other 7 metabolites, i.e. palmitic acid (C_16_H_32_O_2_), stearic acid (C_18_H_36_O_2_), nonadecanoic acid (C_19_H_38_O_2_), eicosanoic acid (C_20_H_40_O_2_), homophytanic acid (C_21_H_42_O_2_), tricosanoic acid (C_23_H_46_O_2_), tetracosanoic acid (C_24_H_48_O_2_), belong to the family of fatty acids. A pathway analysis using MetaboAnalyst 6.0^32^ revealed that palmitic acid, eicosanoic acid and octadecanoic acid play roles in the biosynthesis of unsaturated fatty acids, whereas stearic acid and tetracosanoic acid are involved in the mitochondrial β-oxidation. Notably, all of them showed elevated levels in PD patients, indicating consistent alterations in lipid metabolism, particularly concerning fatty acids. Since some fatty acids like palmitic acid are part of the mitochondrial fatty acid synthesis as well, this observation could be attributed to mitochondrial dysfunction and, thus, defects in mitochondrial metabolism, already associated with PD^38–40^. Recent research has also been suggesting that PD is associated with lipid dysregulation or changes in lipid metabolism^24,41,42^, supporting our findings. Levodopa has been discussed to influence brain fatty acid composition, notably increasing arachidonic acid levels and altering the n-3:n-6 polyunsaturated fatty acid ratio^44^. However, the proposed fatty acid metabolites in this paper have not yet been directly linked to levodopa treatment. Other potential explanations for the observed lipid dysregulation include differences in nutritional intake, variations in microbiome composition, and altered metabolic profiles due to underlying disorders.

Furthermore, dopaminergic drugs used in PD treatment can also contribute to substantial metabolomic differences, particularly in blood plasma^43^. For instance, 3-Methoxytyrosine, likely originating from levodopa, was significantly elevated in the top 15 hits in plasma of PD patients (Supplementary Table 1). As a result, drug-induced metabolites or metabolite patterns may not only cause a misinterpretation in the effective differentiation of both groups but also complicate the identification of endogenous metabolites of interest, as their signals may overlap. Notably, the results pertaining to exhaled breath did not appear to be associated with the dopaminergic medication, as non-manifesting LRRK2 carriers, who were not receiving dopaminergic medication, still exhibited several metabolites that were found to be significantly altered in the PD group (Figure 5, Table 3). This further suggests that the observed metabolomic changes are unlikely to be driven solely by levodopa treatment.

While examining plausible alternatives for all proposed significant metabolite identities within the annotation list, one metabolite was identified as particularly noteworthy. According to HMDB^31^, the chemical formula C_20_H_40_O_2_ annotated as eicosanoic acid can be identified as phytanic acid as well. This branched chain fatty acid has been linked to adult Refsum disease, an autosomal recessive neurological disorder characterized by symptoms like peripheral polyneuropathy, cerebellar ataxia, anosmia, and hearing loss^45,46^. Hence, chronically elevated phytanic acid levels have been reported to be neurotoxic, as it can initiate astrocyte and neural cell death by activating the mitochondrial pathway of apoptosis^47^. Consequently, there is a possibility that phytanic acid could similarly contribute to neuronal cell damage in PD. Furthermore, the metabolite 2-hydroxy-3-methylvaleric acid (HMVA) (C_6_H_12_O_3_) generated by L-isoleucine metabolism, showed lower intensities in PD patients. This compound has been documented to occur at higher levels in the blood and urine of patients suffering from maple syrup urine disease (MSUD)^48^, an inherited metabolic disease predominantly characterized by neurological dysfunction^49^. Branched-chain amino acid alterations have been linked to neurodegenerative movement disorders like PD^50–53^, however, there is currently no established association between HMVA and PD. All the aforementioned results suggest that PD extends beyond being solely a neurodegenerative disease and could be considered a multisystem disorder, affecting various metabolic pathways such as those involving amino acid and fatty acids.

Furthermore, this work not only evaluated potential biomarker candidates for an existing condition but also examined metabolic signatures that may indicate latent disease susceptibility in apparently healthy individuals. Multivariate ROC curve analyses allowed the differentiation of the four healthy individuals carrying a pathogenic variant in *LRRK2* gene from healthy controls based on the metabolomic profiles (Figure 6). In blood plasma, univariate ROC analysis yielded only one common metabolite, the primary fatty acid amide octadecanamide (C_18_H_37_NO), which was significantly lower in both non-manifesting *LRRK2* carriers and PD patients compared to healthy controls (Supplementary Table 1). However, one-way ANOVA showed that these decreased levels were not exclusive to PD patients carrying the *LRRK2* mutation, suggesting that this mutation may not be responsible for its occurrence in PD patients. Nevertheless, as octadecenamide (C_18_H_37_NO) is the amide of stearic acid, this finding supports the idea that PFAMs may play a role in PD. The univariate ROC analysis of exhaled breath revealed interesting results in non-manifesting *LRRK2* carriers. Seven of the 10 top metabolites differentiating PD patients from healthy controls (Figure 5) were also found significantly different (Table 3) and shared the same distribution between the groups. ANOVA results clarified that these metabolites differed significantly between PD patients and healthy controls, irrespective of the *LRRK2* mutation status. This underscores the potential relevance of stearic acid (C_18_H_36_O_2_), eicosanoic acid (C_20_H_40_O_2_), nonadecanoic acid (C_19_H_38_O_2_), nonadecanoyl MG (C_22_H_44_O_4_), palmitic acid (C_16_H_32_O_2_), homophytanic acid (C_21_H_42_O_2_) and HMVA (C_6_H_12_O_3_) in the context of PD, potentially serving as early disease biomarkers (Table 3). Additionally, monoglycerides (MGs) were significantly higher in non-manifesting *LRRK2* carriers (Table 3). An ANOVA confirmed these MGs also differed significantly between healthy individuals and PD patients. Notably, MG levels were not only highest in *LRRK2* patients, but all subgroups exhibited significantly higher levels compared to healthy controls (Figure 7). Moreover, octadecanoyl MG (C_21_H_42_O_4_) and nonadecanoyl MG (C_22_H_44_O_4_) emerged as highly relevant for differentiating PD patients from healthy controls (Supplementary Figure 2), suggesting MGs to be suitable candidates for early disease biomarkers. The exploratory nature of this analysis, however, should be considered. Additionally, the control group showed outliers with elevated values overlapping the range seen in the PD group, as illustrated in the boxplots (Figure 5). This overlap and deviation within the control group raise the possibility that individuals in a prodromal stage of PD may have been inadvertently included among the controls. Future studies incorporating PD biomarkers such as CSF aSyn-SAA will help to elucidate the relationship between prodromal PD and early breathome changes.

Considering the presence of pathogenic variants within the PD patient group, this study presents specific metabolite patterns that enable the distinction of the subgroups (Supplementary Figures 5-7). However, the classification of the subgroups based on metabolomic profiles in both body fluids was only partially accurate (Supplementary Figure 7). Especially patients with pathogenic variants in *LRRK2* and *PRKN* genes were misclassified, whereas those with a *GBA1* variant showed better results for both body fluids. This finding implies that *GBA1* mutations potentially lead to more distinctive metabolic changes compared to the other mutations. However, small sample sizes (12 with *LRRK2* and 6 with *PRNK* mutation) limit the results (Table 1A). Nevertheless, discernible differences exist between PD patients with and without pathogenic variants, as all IPD patients were correctly classified based on the metabolic pattern compared to patients with pathogenic variants (Supplementary Figure 7). However, the small number of discernible metabolites identified in a volcano analysis (10 in exhaled breath and 23 in blood plasma analysis) (Supplementary Figures 5, 6) indicates that, as expected based on the many similarities between IPD and genetic PD, the distinctions are not as pronounced as when differentiating between PD patients and healthy controls, wherein the volcano analysis identified over 80 metabolites.

While our study provides valuable insights into the metabolic profile in PD patients, certain limitations should be acknowledged to ensure accurate interpretation. The biggest challenge of this untargeted approach using high-resolution MS remains assigning identified masse to molecular structures^54^. However, while the proposed identities are putative, the indicated chemical formulas are accurate (mass error < 1 ppm). Limitations pertaining to our methodological approach have been addressed in our previous paper^30^. Furthermore, it should be considered that potential comorbidities in patients that could have an influence on the metabolome were not taken into account. Another limitation of the study design is the inclusion of two independent control groups for breath and plasma analysis. However, the independent control groups strengthened the validity of breath as a promising alternative biofluid, as metabolite overlaps between PD patients and controls were observed in both fluids. One additional constraint is the varied PD stages among patients, making it challenging to correlate metabolite profiles with PD progression. A more homogeneous patient population and additional clinical data are required to correlate metabolite patterns with symptom severity. Lastly, the small sample size, particularly of only four non-manifesting carriers, necessitates larger metabolomics studies to validate the proposed biomarker candidates. Conducting longitudinal studies is crucial for tracking individual metabolomic changes and determining if they later develop symptomatic conditions.

In conclusion, metabolomic profiles of blood plasma and exhaled breath effectively differentiated PD patients from healthy controls, revealing distinct metabolic patterns. Substantial differences were observed between these fluids, indicating that alternative matrices like breath add valuable information about (patho)physiological processes. Our untargeted metabolomic approach delineates a non-volatile metabolome of PD, laying the foundation for assessing clinical biomarkers in exhaled breath. The metabolomic breath profiling identified 10 significant metabolites. Seven of these were also detected in non-manifesting *LRRK2* carriers, suggesting a potential link to early PD development before clinical symptoms appear. Most of the proposed metabolites are intermediates in fatty acid metabolism, introducing new hits for breath analysis in PD. Future investigations should confirm their identities and assess their role in PD pathophysiology and progression, as well as their utility as biomarkers for early diagnosis. Nonetheless, evaluating differences in the breathome of PD patients marks the initial stride towards establishing a diagnostic breathprint, eventually paving the way for targeted and effective interventions.

## METHODS

### Study design

By employing a previously established method^30^, this experimental study performs metabolomic profiling of exhaled breath of PD patients, both with and without pathogenic variants. Between July 2021 and August 2023, this project enrolled patients who were genetically characterized in either the LIPAD^55^ or ROPAD^6^ study, or both. The inclusion criteria required PD diagnosis according to the Movement Disorder Society (MDS) and participants aged at least 18^6,55^.

Additionally, the study population was genetically characterized based on the presence of pathogenic variants. PD patients were subdivided into a group without a positive genetic testing report (idiopathic PD) and patients with genetic variants in the *LRRK2*, *PRKN*, and *GBA1* genes. The *LRRK2* group comprised participants harboring one of the following *LRRK2* variants: p.Gly2019Ser (c.6055G>A), p.Arg1441Cys (c.4321C>T), p.Tyr1699Cys (c.5096A>G)^55^.

Furthermore, to evaluate the potential of breath analysis, we used the commonly used diagnostic body fluid blood plasma for comparison. Rather than directly comparing breath and blood plasma samples from the same individual, our objective was to assess the overall utility of breath analysis relative to blood sample analysis. To ensure an independent evaluation of the diseased sample groups, we included two distinct healthy control groups: one providing breath samples and the other providing blood plasma samples. Both groups consisted of healthy individuals without a PD diagnosis.

Additionally, healthy individuals with a first- or second-degree relative who tested positive for a genetic variant (*LRRK2*, *GBA1*) were also recruited. The inclusion criteria for the healthy breath control group were consistent with previous reports^30^

This study adhered to the principles outlined in the Declaration of Helsinki (as revised in 2013) and received ethical approval from the Ethics Committee of the University of Lübeck, Germany. Prior to participation, all patients gave informed consent. The methodological scheme of the study is presented in Supplementary Figure 8.

### Specimen collection and Sample Preparation

#### Exhaled Breath

Exhaled breath samples were collected non-invasively utilizing a device equipped with a polymeric electret filter (three replicates per participant). The sample acquisition and extraction of the electret filter was performed according to established protocols^30,56^. For mass analysis, the residue from each sample was re-suspended in 350 µl methanol/H_2_O (1:1, v/v) and subsequently diluted 1:20 with methanol/H_2_O (1:1 v/v) in 1.5 mL HPLC vials.

#### Blood Plasma

Plasma extraction from blood samples involved the collection of blood into two EDTA tubes, ensuring proper filling to maintain sample integrity. Following collection, each tube was carefully inverted ten times to ensure thorough mixing. Subsequently, the tubes were immediately placed on ice to prevent protein degradation. The sample tubes were centrifuged at 2000 g for 10 minutes at a temperature of 4°C. After centrifugation, the resulting supernatant was transferred into a new 15 ml sterile polypropylene falcon tube and inverted to ensure homogeneity. The supernatant was then aliquoted into twelve 300 µl Cryovials, with each aliquot promptly frozen at -80°C to preserve sample quality until further analysis.

Further sample preparation used 500 µL of the blood plasma. The plasma samples were extracted using a modified approach^57^, ensuring the efficient isolation and preparation of each sample component for further mass analysis. Extraction of the 500 µL blood samples resulted in a lipophilic and hydrophilic phase, as well as a protein pellet.

First, 500 µL methanol and 4 ml methyl *tert*-butyl ether (MTBE) were added to 500 µL of sample and vortexed. Samples were incubated at room temperature for 30 minutes using an overhead shaker with 25 rounds per minute (Trayster, Ika, Staufen, Germany). In the next step, 500 µL of ultra-pure water was added, and the mixture was then incubated for 10 minutes at room temperature without agitation. Lastly, all samples underwent centrifugation (Allegra X-30R/SX 4400, Beckman Coulter, Krefeld, Germany) at 1,000 *g* for 10 minutes.

The lipophilic phase was carefully transferred to a fresh 15 mL tube and stored at 4°C for subsequent use. A second extraction cycle was conducted using the hydrophilic phase, following the same extraction steps as outlined before. Upon completion of the second extraction cycle, the lipophilic phases were combined and the hydrophilic phase was carefully transferred into a separate 15 mL tube, while the protein pellet was discarded.

The hydrophilic and lipophilic phases were dried using a vacuum concentrator (SpeedVac, thermo fisher scientific, Bremen, Germany) and re-suspended, respectively. Thus, the hydrophilic and lipophilic phases were diluted at 1:1000 in the solvents methanol/H_2_O (1:1, v/v) and isopropanol/chloroform (3:1, v/v), respectively. The vials were stored at -80 °C until analysis *via* mass spectrometry.

#### FT-ICR-MS measurements

This study utilized an ultra-high-resolution FT-ICR-MS system (7 Tesla, Solari-XR-X, Bruker, Bremen, Germany) coupled with an Infinity 1260 HPLC for direct sample injection (Agilent, Waldbronn, Germany). The mass spectrometric analysis was conducted according to our established method with identical parameters^30^.

Electrospray ionization was used in both positive and negative modes. Two different measurement methods detected ultra-small molecules and small molecules achieving a detection range of 65-1500 m/z. The FT-ICR-MS featured a 2-omega cell with a precision of attained m/z signals to less than 1 ppm. The ICR-cell was calibrated utilizing sodium trifluoroacetate to achieve an accuracy of < 0.5 ppm. Quality control samples, pooled from all samples, were prepared to assess sample stability and dilution^30,58^.

#### Sample evaluation and data processing

The raw data was processed using the MetaboScape 2021b software (Bruker, Bremen, Germany). After importing, the dataset was recalibrated (accuracy <1 ppm) with a calibration list, containing > 60 diverse matrix metabolites such as amino acids, carbohydrates or various lipids. The HMDB^31^ 2023 was used for metabolite annotation. Metabolites not included in the database were not identified in the dataset. In order to consolidate the dataset, further analysis steps comprised data normalization, duplicate removal and data reduction of replicates.

First, the calculated bucket tables were exported to R 4.3 and all data was normalized using a PQN normalization script^59^. Consequently, the normalized data originating from the two measurement techniques as well as from both ionization modes were merged to create one single dataset covering the complete mass range (65-1500 m/z). Second, a method for duplicate removal was applied^59^. Accordingly, duplicate metabolites identified in both ionization techniques were eliminated by selecting the feature with the highest overall intensity across all samples for further analysis. Duplicates with lower scores were subsequently removed. The third and last step involved data reduction of the three replicates. To be qualified, a metabolite needed to be present in at least two of the three replicates obtained from the participants. Subsequently, the mean intensities of the replicates were calculated. In cases where a metabolite was detected in only one sample out of the three replicates, it was designated as “not detected” for that particular participant. Finally, the acquired dataset was used to delineate the non-volatile core metabolome of PD patients with and without pathogenic variants. Further, biostatistical analyses were applied to identify significant metabolites.

#### Statistical Analyses

The processed and evaluated data underwent biostatistical analyses using MetaboAnalyst 6.0^32^ with peak intensities and samples organized in columns (unpaired). A fixed value of 1/5 of the limit of detection was used to address the missing (zero) values. To identify the most stable subgroup-specific metabolites and reduce the potential impact of individual variations, further data processing and statistical analyses focused on metabolites detected in at least 10% of all samples. Statistical analysis applied median intensity values and the data used median normalization, log transformation, and Pareto scaling for normal distribution.

The metabolome analyses were performed employing metabolite set enrichment analysis (MSEA) with 1250 sets of metabolite-sub-classes. Metabolites present exclusively in healthy individuals but absent in PD patients were excluded prior to the MSEA to enhance the accuracy of our comparisons. Furthermore, univariate and multivariate analyses were combined to discover relevant metabolites in both breath and blood plasma. To compare two groups such as PD patients and healthy controls, we performed a volcano analysis. The fold-change was set to >1 (double effect size between the groups). The adjusted *p*-value was set to < 0.1.

To enable comparisons between multiple groups such as PD patients with different pathogenic variants, we applied a multiple ANOVA combined with a Tukey honestly significant difference (HSD) test for false discovery rate (FDR) correction. The significance threshold was set at a *p*-value of < 0.05. In addition, a partial least squares discriminant analysis (PLS-DA) was performed to generate scores plot. This analysis identified the variable importance for projection (VIP) features, highlighting pattern differences between various subgroups.

Moreover, a classification analysis was conducted employing a random forest algorithm, integrated within the MetaboAnalyst 6.0^32^ software. The algorithm automatically picked random samples to generate a training-dataset. Subsequently, a separate test-dataset from the uploaded data was used to determine whether classification could be successfully performed based solely on metabolites. The seeds were selected randomly. Additionally, a maximum of 500 decision trees were permitted. The classification of two distinct subgroups was determined by both the out-of-bag (OOB) error and the classification error (class.error).

The significant metabolites obtained from the volcano analysis, PLS-DA and the random forest classification were combined in a Venn diagram, thereby evaluating the applicability of each test for analyzing the respective body fluid and identifying the most reliable metabolites across all three statistical approaches.

To evaluate the diagnostic power of particular metabolites, univariate and multivariate exploratory Receiver Operating Characteristic (ROC) analysis were executed with MetaboAnalyst Biomarker Analysis 6.0^32^. Herein, the PLS-DA and its built-in feature were selected for the classification and ranking methods, respectively. The analysis was conducted using two latent variables.

The ROC curves were generated applying Monte-Carlo cross-validation (MCCV) with balanced sub-sampling, e.g. an equal number of positive and negative cases. Specifically, each MCCV iteration involved randomly splitting the dataset into two parts. The training set used two thirds of the samples to elaborate on the importance of the feature. The most important metabolite features (max. top 100) were subsequently used to build classification models. These models were validated on the remaining one-third of the samples (testing set). This process was repeated multiple times to calculate the performance and confidence interval of each model. Furthermore, classical univariate ROC analysis for single biomarkers used 500 bootstrappings for calculating the 95% confidence interval.

## Data Availability

All data produced in the present study are available upon reasonable request to the authors

## ACKNOWLEDGMENTS

The authors sincerely thank all volunteers and patients for participating in this study.

## AUTHOR CONTRIBUTIONS

*M.M.* designed and conceptualized the study, took breath samples from the participants, collected and prepared all breath and plasma samples, carried out the experiments, prepared the figures, performed biostatistical analysis, and wrote the manuscript. *T.U*. and *M.B*. were involved in recruiting patients and collecting samples. *T.D.* carried out the MS analysis, metabolite annotation and supported in performing biostatistical analysis. *K.S.* was involved in interpreting the data. *K.L.* was involved in genetic testing and supervising preparation of blood plasma samples. . *P.B.*, *N.B.* and *C.K.* were involved in recruiting patients, designing and conceptualizing the study. *T.K.* conceived, designed, and supervised the metabolomics part of the study. C.K. and T.K. jointly supervised this work and were involved in manuscript writing.

All authors revised and approved the manuscript. Correspondence and request for materials should be addressed to M. Malik, C. Klein or T. Kunze.

