## Supplementary Information for "Metabolomic breath landscape analysis unravels lipid biomarker candidates in patients with monogenic and idiopathic Parkinson’s disease"

**Supplementary Figure 1**

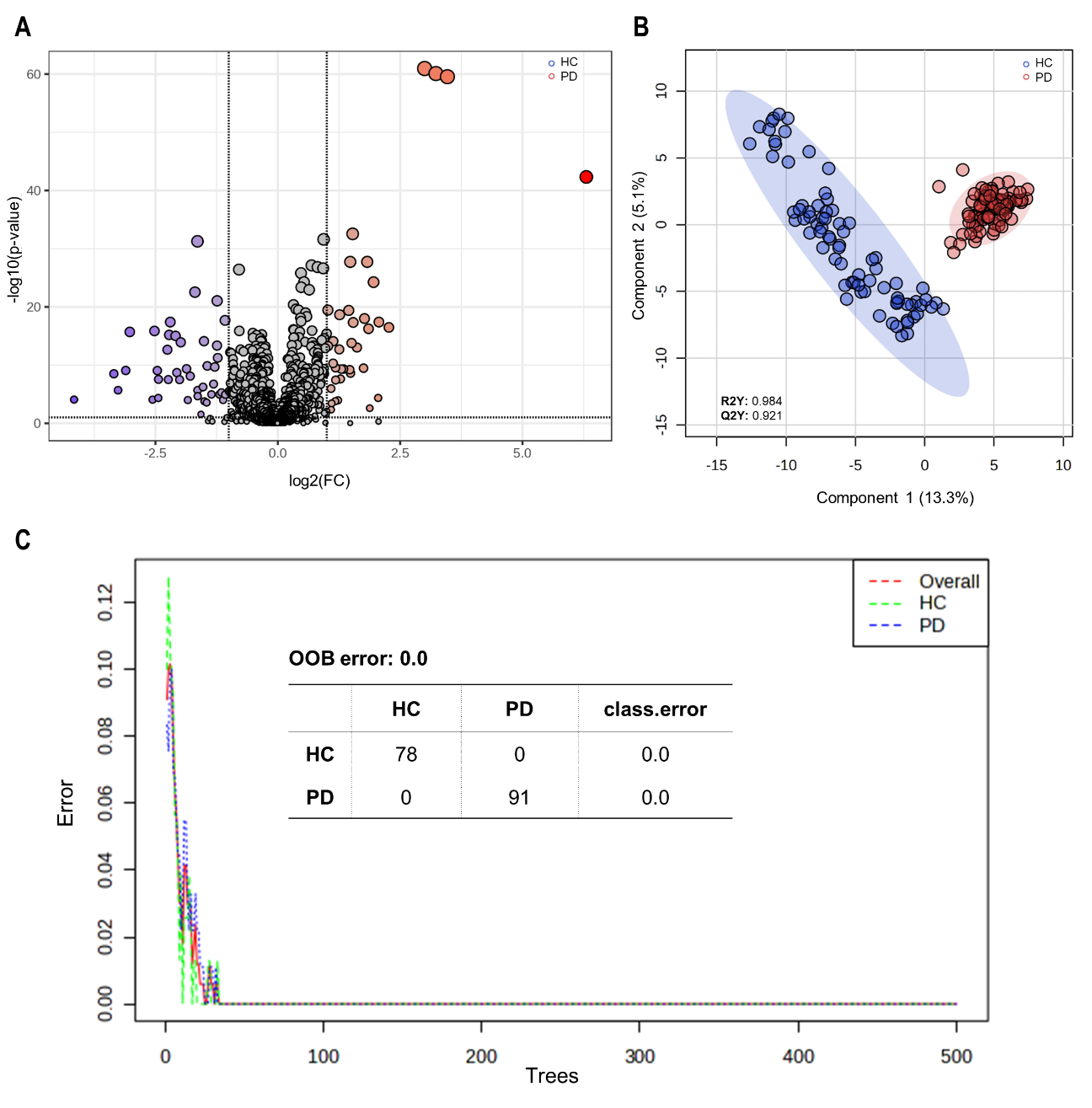

**Supplementary Figure 1: Elaboration of metabolic patterns in blood plasma using three biostatistical methods. A** Univariate analysis: 82 metabolites identified by volcano plot differing between PD patients and healthy controls (HC), meeting the criteria of a fold change (FC) threshold > 2.0 and an adjusted p-value (FDR correction) of < 0.1. Metabolites exhibiting a log2(FC)> 0 (red) indicate elevated intensities in PD patients, while those with a log2(FC) < 0 (blue) signify higher levels in HC. **B** Multivariate analysis: Scores plot of PLS-DA analysis depicting a clear separation of the metabolite profile of PD patients (red) and HC (blue) with R2Y= 0.984 and Q2Y= 0.921 for five components. **C** Classification analysis: Random forest classification with 46 predictors and 500 trees. Based on their metabolite profile, all PD patients as well as all HC were correctly classified. All calculations and analyses were carried out using MetaboAnalyst 6.0.

**Supplementary Figure 2**

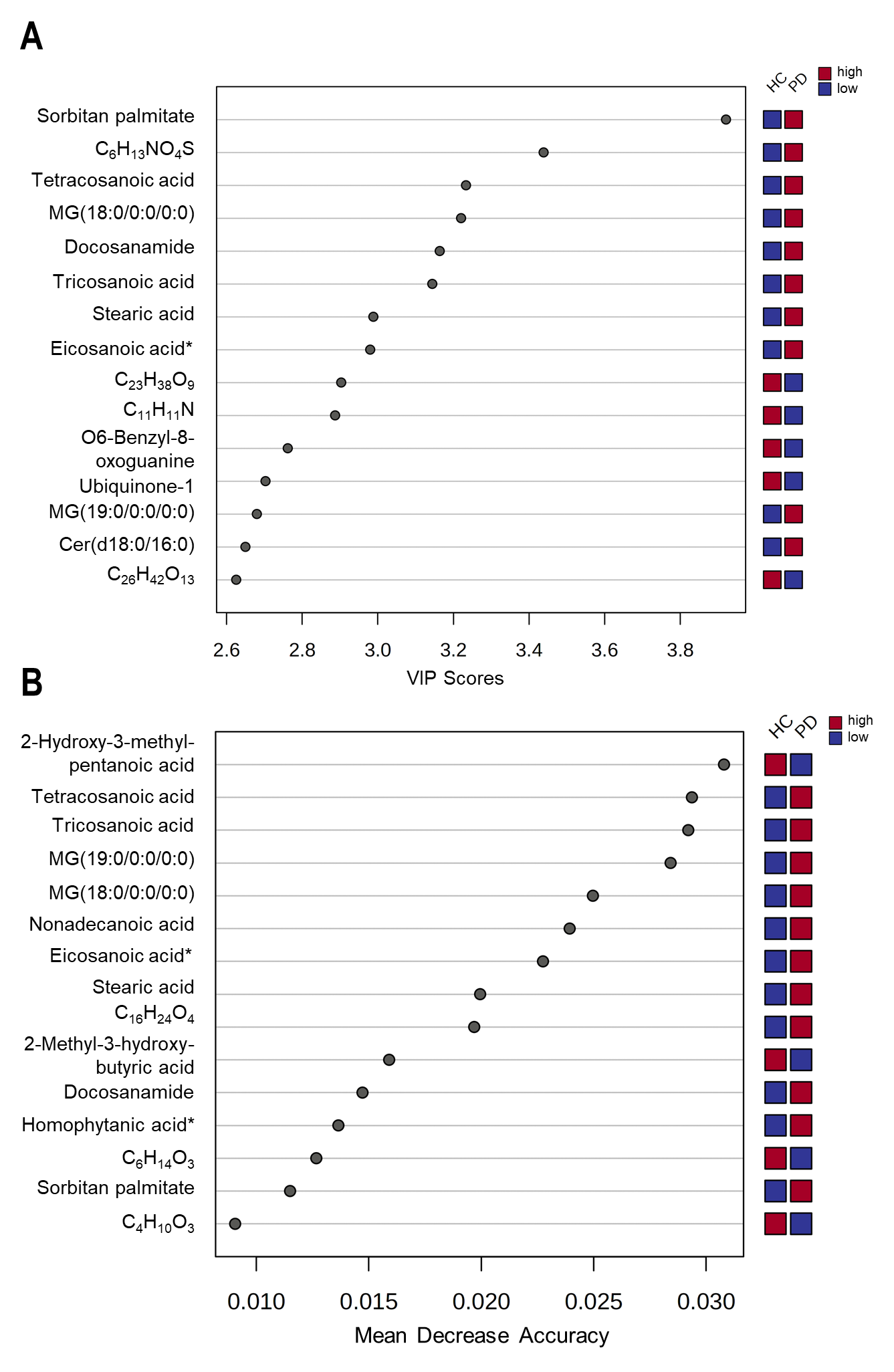

**Supplementary Figure 2: The top 15 metabolites in exhaled breath identified by PLS-DA and random forest analysis. A** The top 15 VIP features (VIP Scores > 2.5) are presented, with the coloured boxes on the right indicating the relative abundance (high: red, low: blue) of the corresponding metabolite in the group of PD patients compared to healthy controls (HC). **B** The top 15 features identified in the random forest analysis (40 predictors, 500 trees) with a mean decrease accuracy > 0.001. The coloured boxes on the right indicate the relative abundance (high: red, low: blue) of the corresponding metabolite in the group of PD patients compared to HC. All calculations and analyses were carried out using MetaboAnalyst 6.0. In cases, the metabolite annotation was not plausible, only the chemical formula is provided. Putative metabolites were labelled with an asterisk (*) when metabolite annotation assigned two distinct metabolites for one chemical formula. The chemical formula annotated as eicosanoic acid and homophytanic acid can also represent phytanic acid and heneicosanoic acid, respectively.

**Supplementary Figure 3**

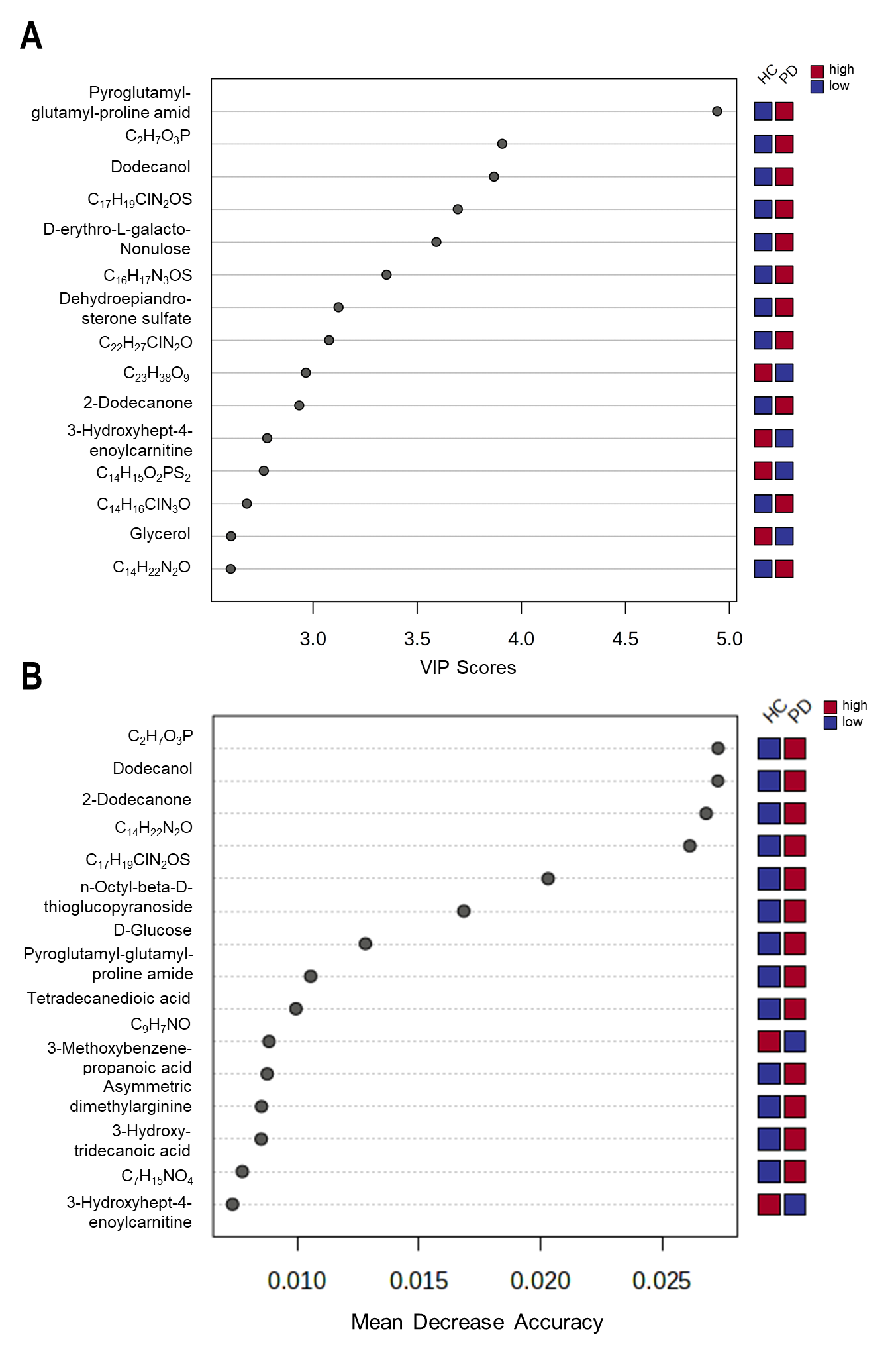

**Supplementary Figure 3: The top 15 metabolites in blood plasma identified by PLS-DA and random forest analysis. A** The top 15 VIP features (VIP Scores > 2.5) are presented, with the coloured boxes on the right indicating the relative abundance of the corresponding metabolite (high: red, low: blue) in the group of PD patients compared to healthy controls (HC). **B** The top 15 features identified in the random forest analysis (46 predictors, 500 trees) with a mean decrease accuracy > 0.001. The coloured boxes on the right indicate the relative abundance (high: red, low: blue) of the corresponding metabolite in the group of PD patients compared to HC. All calculations and analyses were carried out using MetaboAnalyst 6.0. In cases, the metabolite annotation was not plausible, only the chemical formula is provided.

**Supplementary Figure 4**

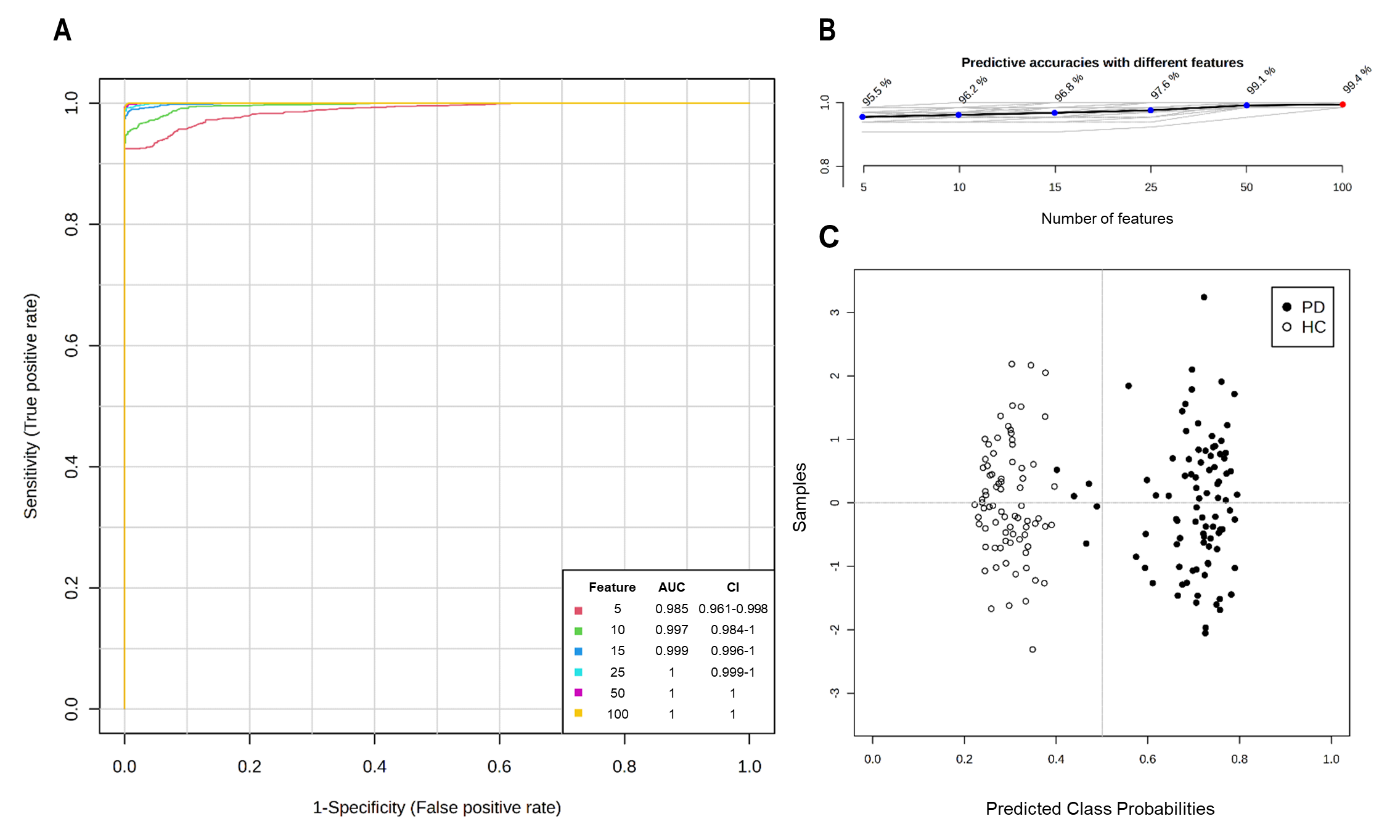

**Supplementary Figure 4: Data-based PD prediction via multivariate exploratory ROC curve analysis of blood plasma samples.** PLS-DA and its built-in feature was selected as the classification method and the ranking method, respectively. The analysis was conducted using two latent variables. **A** ROC curve analyses for n = 5, 10, 15, 25, 50 and 100 independent metabolite features in PD patients and healthy controls (HC). AUC and 95% confidence intervals (CI) were calculated by Monte Carlo cross validation (MCCV) using balanced sub-sampling **B** Predictive accuracies (y-axis) in percent (%) for n = 5, 10, 15, 25, 50 and 100 independent metabolite features. **C** Predicted class probabilities using the best classifier (n=25) based on the AUC. Using this model, all healthy controls (n=78) were accurately predicted while five of 91 PD patients were classified as healthy false negatively. Since a balanced sub-sampling technique was implemented during model training, the classification boundary consistently aligns with x = 0.5, indicated by the dotted line. ROC curve analyses were carried out using MetaboAnalyst Biomarker Analysis 6.0.

**Supplementary Figure 5**

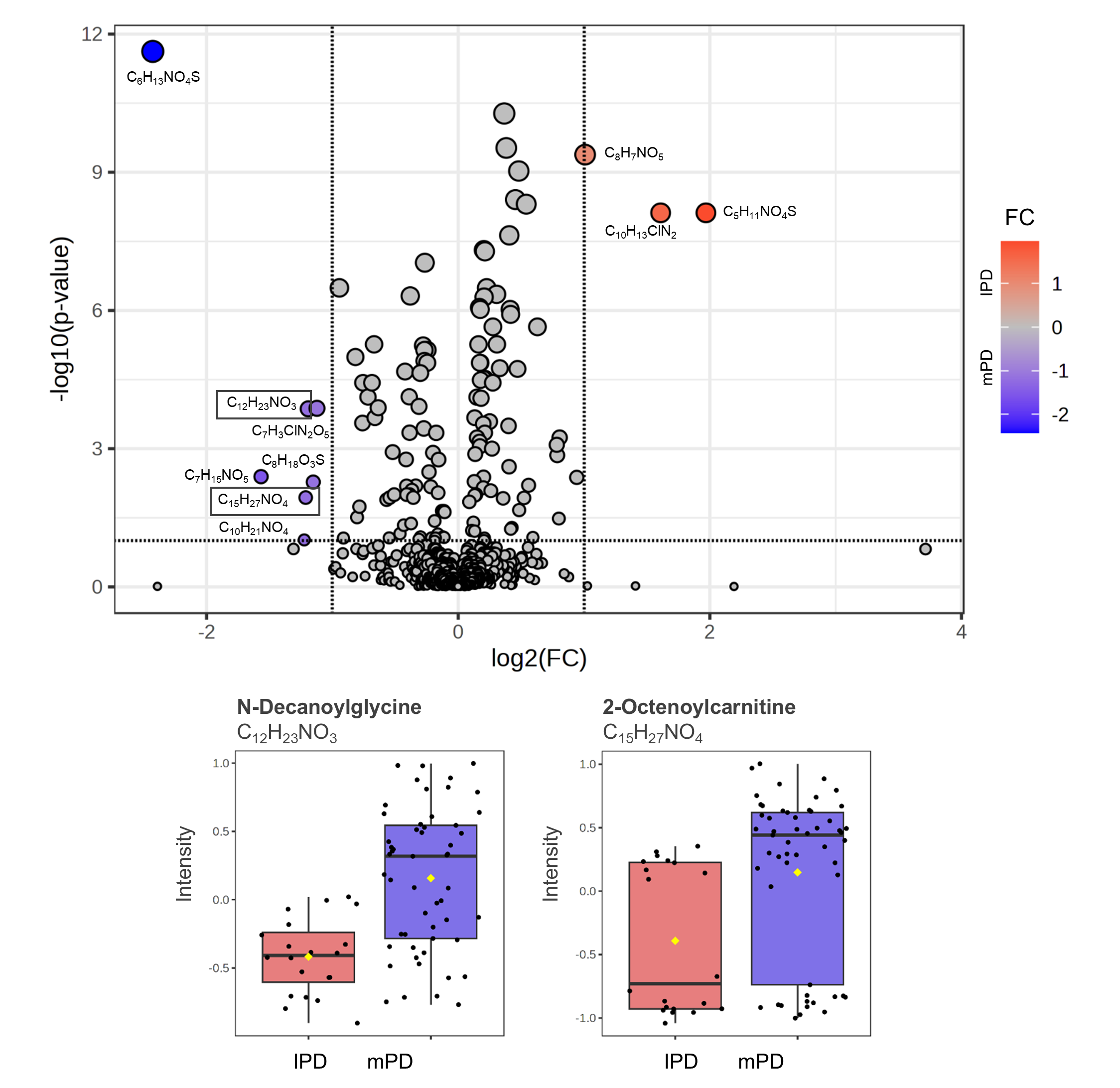

**Supplementary Figure 5: Volcano analysis of IPD patients vs. PD patients with a mutation (mPD).** Volcano plot with a FC threshold > 2.0 and an adjusted p-value (FDR correction) of < 0.1 identified 10 metabolites differing between IPD patients and PD patients carrying a mutation in GBA, LRRK2 or PRKN genes (mPD). Metabolites exhibiting a log2(FC)> 0 (red dots) indicate elevated intensities in IPD patients, while those with a log2(FC) < 0 (blue dots) signify higher levels in patients carrying one of the aforementioned mutations (mPD). The red and blue dots represent a single metabolite, each labelled with its chemical formula. Relevant and endogenously occurring metabolites are depicted using box plots comparing IPD patients (red, n= 20 biologically independent exhaled breath samples) and mPD patients (blue, n= 53 biologically independent exhaled breath samples). Black dots represent the values from all samples. The box and whiskers summarise the normalised values with the centre line presenting the median. The mean value is indicated as a yellow diamond.

**Supplementary Figure 6**

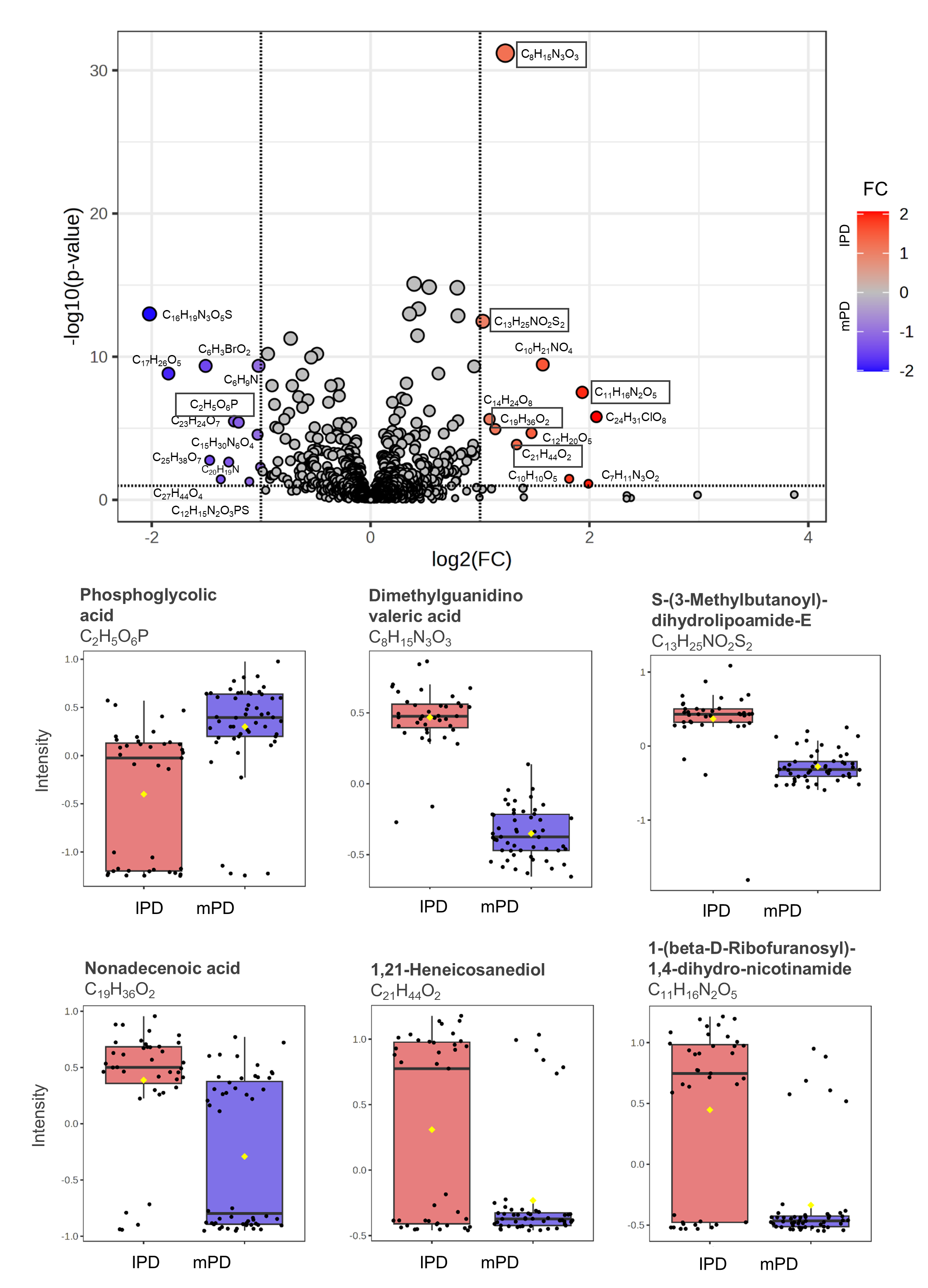

**Supplementary Figure 6: Volcano analysis of IPD patients vs. PD patients with a mutation (mPD).** Volcano plot with a FC threshold > 2.0 and an adjusted p-value (FDR correction) of < 0.1 identified 23 metabolites differing between IPD patients and PD patients carrying a mutation in GBA, LRRK2 or PRKN genes (mPD). Metabolites exhibiting a log2(FC)> 0 (red dots) indicate elevated intensities in IPD patients, while those with a log2(FC) < 0 (blue dots) signify higher levels in patients carrying one of the aforementioned mutations (mPD). The red and blue dots represent a single metabolite, each labelled with its chemical formula. Relevant and endogenously occurring metabolites are depicted using box plots comparing IPD patients (red, n= 39 biologically independent exhaled breath samples) and mPD patients (blue, n= 52 biologically independent exhaled breath samples). Black dots represent the values from all samples. The box and whiskers summarise the normalised values with the centre line presenting the median. The mean value is indicated as a yellow diamond.

**Supplementary Figure 7**

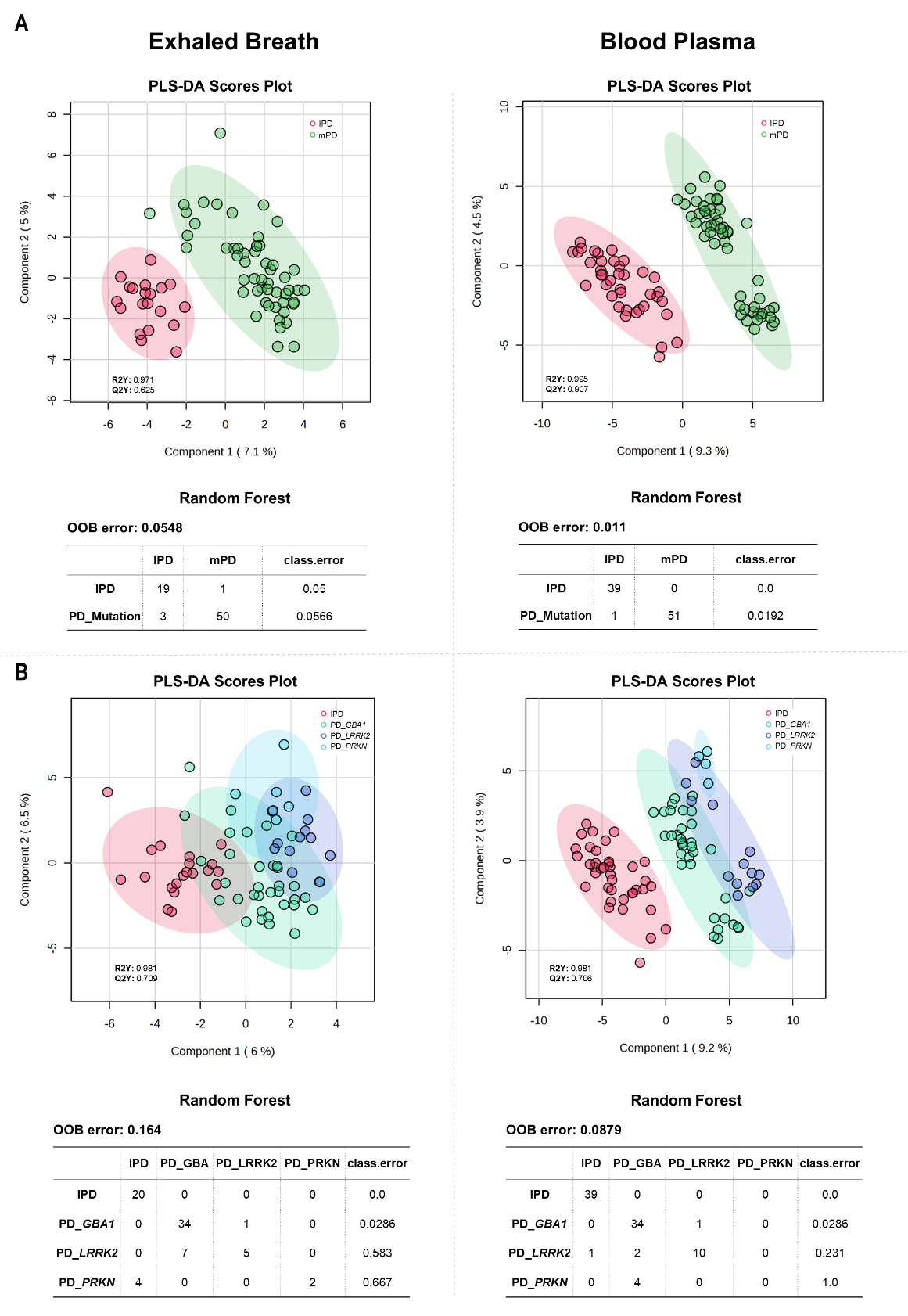

**Supplementary Figure 7: PD subgroup analysis of exhaled breath and blood plasma using PLS-DA and random forest analysis. A** Scores plot of PLS-DA analysis for exhaled breath (left) and blood plasma (right) depicting a separation of the metabolite profiles of IPD patients (red) and PD patients carrying a mutation (mPD = GBA, LRRK2 or PRKN) (green). Random forest classification was performed with 500 trees and 40 predictors, resulting in an OOB error of 0.055 and 0.011 for exhaled breath and blood plasma, respectively. **B** Scores plot of PLS-DA analysis for exhaled breath (left) and blood plasma (right) presenting overlapping and distinctive clusters of the metabolite profiles of the PD patients: (1) IPD (red) and subgroups of PD patients carrying a specific mutation (2) PD_GBA (green), (3) PD_LRRK2 (blue) and (4) PD_PRKN (turquoise). Random forest classification was performed with 500 trees and 40 predictors, resulting in an OOB error of 0.164 and 0.088 for exhaled breath and blood plasma, respectively.

**Supplementary Figure 8**

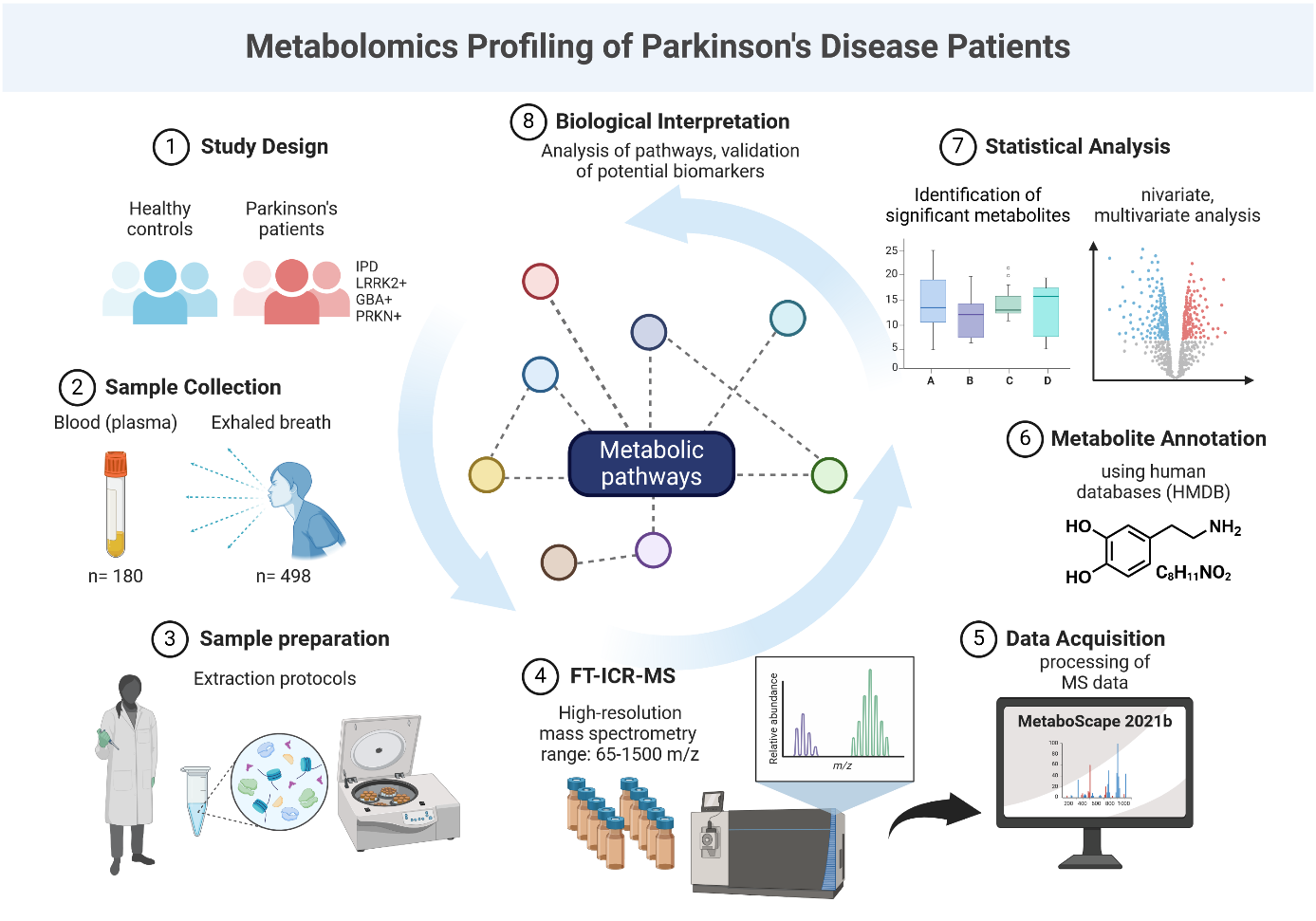

**Supplementary Figure 8: Methodological scheme of the study.** The study population comprised idiopathic and monogenic PD patients (GBA, LRRK2 and PRKN) as well as healthy controls. Exhaled breath samples (n= 498) and blood plasma samples (n= 180) were collected and analysed utilising high-resolution FT-ICR-MS. The mass data was processed in MetaboScape 2021b and metabolite annotation was conducted in reference to the human metabolome database (HMDB). Further data processing included biostatistical analyses that were performed using MetaboAnalyst 6.0. Metabolomic profiles of PD patients and healthy individuals were compared to uncover potential biomarker candidates for PD. This figure was created with BioRender (biorender.com).

**Supplementary Table 1**

**Supplementary Table 1:** **The 15 most relevant metabolites in blood plasma of PD patients obtained from Venn analysis.** The metabolites were identified in all three biostatistical methods (PLS‑DA, volcano plot, random forest) and then subjected to Venn diagram analysis. This table characterizes the most robust metabolites by expression ratios obtained from a fold change analysis (FC=2.0), adjusted p‑values <<0.001, VIP scores > 1.5 and mean decrease accuracy values >0.0001, with arrows indicating either higher or lower levels in PD patients compared to healthy controls. In cases the metabolite annotation was not plausible, only the chemical formula is provided. Metabolites labelled with a hash mark (#) were identified in Venn analysis of exhaled breath as well.

| Chemical formula | Putative identity | Expression ratio (FC) | Volcano plot (adj. p‑value) | PLS-DA (VIP Score) | Random forest (MDA) |
| --- | --- | --- | --- | --- | --- |
| C12H26O | Dodecanol | ↑ 0.11 | 5.89E-61 | 3.94 | 0.024 |
| C2H7O3P | Ethylphosphonic acid | ↑ 0.09 | 2.06E-60 | 3.98 | 0.023 |
| C14H28O5S | n-Octyl-beta-D-thioglucopyranoside | ↑ 0.35 | 2.36E-33 | 2.55 | 0.016 |
| C9H7NO | Indole-3-carboxaldehyde# | ↓ 3.12 | 6.51E-32 | 3.03 | 0.007 |
| C12H24O | 2-Dodecanone | ↑ 0.28 | 1.80E-28 | 2.99 | 0.021 |
| C14H25NO5 | 3-Hydroxyhept-4-enoylcarnitine | ↓ 3.21 | 3.31E-23 | 2.84 | 0.007 |
| C14H22N2O | unknown identity# | ↑ 0.36 | 1.71E-28 | 3.14 | 0.029 |
| C9H18O9 | D-erythro-L-galacto-Nonulose | ↑ 0.26 | 6.47E-25 | 3.66 | 0.006 |
| C16H29NO5 | 6-Hydroxynon-2-enoylcarnitine | ↑ 0.49 | 3.91E-20 | 2.17 | 0.001 |
| C7H15NO4 | unknown identity | ↑ 0.36 | 4.26E-20 | 2.52 | 0.004 |
| C18H37NO | Octadecanamide | ↓ 2.35 | 8.358E-22 | 1.96 | 0.004 |
| C19H28O5S | Dehydroepiandro-sterone sulfate | ↑ 0.29 | 1.03E-18 | 3.18 | 0.002 |
| C8H18N4O2 | (A)symmetric dimethylarginin | ↑ 0.34 | 4.77E-18 | 2.51 | 0.009 |
| C9H17NO3 | N-Heptanoylglycine | ↓ 5.70 | 1.33E-16 | 2.33 | 0.010 |
| C10H13NO4 | 3-Methoxytyrosine# | ↑ 0.35 | 1.58E-14 | 2.24 | 0.004 |
